# Daith piercing, a social media hype on YouTube for the treatment of migraine? A systematic video analysis of quality and reliability

**DOI:** 10.1101/2025.01.19.25320657

**Authors:** Saroj K. Pradhan, Michael Furian, Giada Todeschini, Qiong Schürer, Xiaying Wang, Bingjun Chen, Yiming Li, Andreas R. Gantenbein

## Abstract

**Background:** Nowadays, social media video-sharing website YouTube is globally accessible and used for sharing news and information. It also serves as a tool for migraine sufferers seeking guidance about Daith piercing as a potential migraine treatment; however, shared and disseminated video content is rarely regulated and does not follow evidence-based medicine.

**Objective:** To investigate the content, quality, and reliability of YouTube videos on Daith piercing for the treatment of migraine.

**Methods:** YouTube videos were systematically searched from the video portal inception until 17^th^ January 2024. “Daith piercing” AND “migraine” were the applied search terms. All video blogs reporting on Daith piercing for the treatment of migraine were included. The primary outcome of interest was assessing the Global Quality Scale and DISCERN implementing the five-point Likert scale (“very poor” to “excellent”) to evaluate each video blog’s quality, flow, and reliability. Secondary outcomes included the relapse time of migraine after Daith piercing, and further outcomes related to Daith piercing. This systematic video analysis was preregistered in PROSPERO (https://www.crd.york.ac.uk/prospero/ CRD42024510089).

**Results:** Out of 496 initially screened video recordings, 246 videos were included in the final analysis (N=69 categorized as *Personal Experience*; N=176 as *Others,* defined as videos from bloogers, piercers or other persons not affected by migraine; and N=1 as *Healthcare Professionals*). Overall, these videos collectively gathered 67783 likes and received 4.68 million views. The Global Quality Scale rating in the category *Personal Experience* revealed that the quality of 50.7% of videos was very poor; 29.0% poor; 11.6% moderate and 8.7% good. In the category *Others*, Global Quality Scale rating showed that the quality of 60.8% of videos was very poor; 25.6% poor; 11.9% moderate and 1.7% good (P<0.05 compared to the proportion of good quality videos in the category *Personal Experience* quality). The one video in the category *Healthcare Professionals* was rated “poor quality”. Ratings applying the DISCERN tool were comparable. Overall, 111 (45.1%) videos recommended and 14 (5.7%) discouraged Daith piercing for migraine relief.

**Conclusion:** Based on the Global Quality Scale and DISCERN scores, the information, usefulness, and accuracy of most YouTube content on Daith Piercing for migraine treatment are generally of poor quality and reliability. The lack of high-quality and reliable videos might expose users to potentially misleading information and the dissemination of unproven medical interventions.

**Plain Language Summary:** This systematic video analysis evaluated the quality and reliability of 246 YouTube videos on Daith piercing as a migraine treatment, accessing the DISCERN instrument and the Global Quality Scale tool. The analysis revealed that most videos were anecdotal, lacked input from healthcare professionals, and demonstrated poor quality and reliability. These findings highlight that YouTube is not a reliable source for medical information on Daith piercing for migraine treatment, underscoring the importance of consulting physicians and referencing peer-reviewed research for well-informed decision-making.

## Introduction

Migraine is a complex, recurring neurobiological disorder that is still not fully understood in its pathophysiology.^1^ Daith piercing (DP) also known as the “migraine piercing”, consists in piercing the crus of the helix (Figure 1) and has gained popularity in recent years. Numerous video blogs on social media, such as YouTube, assert that DP serves as an alternative treatment for migraine, and anecdotal patient reports on the internet suggest an improvement in migraine symptoms. In contrast, certain headache societies have released statements not supporting the use of DP as a treatment for migraine.^2,3^ A recent literature search about DP in migraine treatment discovered only one narrative review^4^ and no clinical trials. The available literature, along with the documented recurrence of pain and the accompanying side effects attributed to DP, suggest that the current evidence does not substantiate the use of DP as an effective treatment for migraine.^4^ Moreover, we found insufficient evidence for DP against migraine-related pain relief.^4^ As individuals with chronic or recurrent migraines progressively turn to the World Wide Web to obtain health-related facts, the accessibility of a vast array of such information has become notably expedient. The use of YouTube for health-related information has witnessed a major and post content, facilitating easy communication and commentary among users.^5^ However, leveraging the web for health information presents various drawbacks, particularly the intricacy of medical terminology, the scarcity of structure, and the absence of evidence-based medicine and regulation. Consequently, understanding the accuracy and reliability of disseminated information is crucial. Due to the absence of evidence but increasing popularity of applying health-related measures promoted on YouTube, the primary aim of this systematic video analysis was to evaluate the current state of quality and reliability of published YouTube videos concerning the efficacy of DP in relieving migraine.

**Figure 1:**
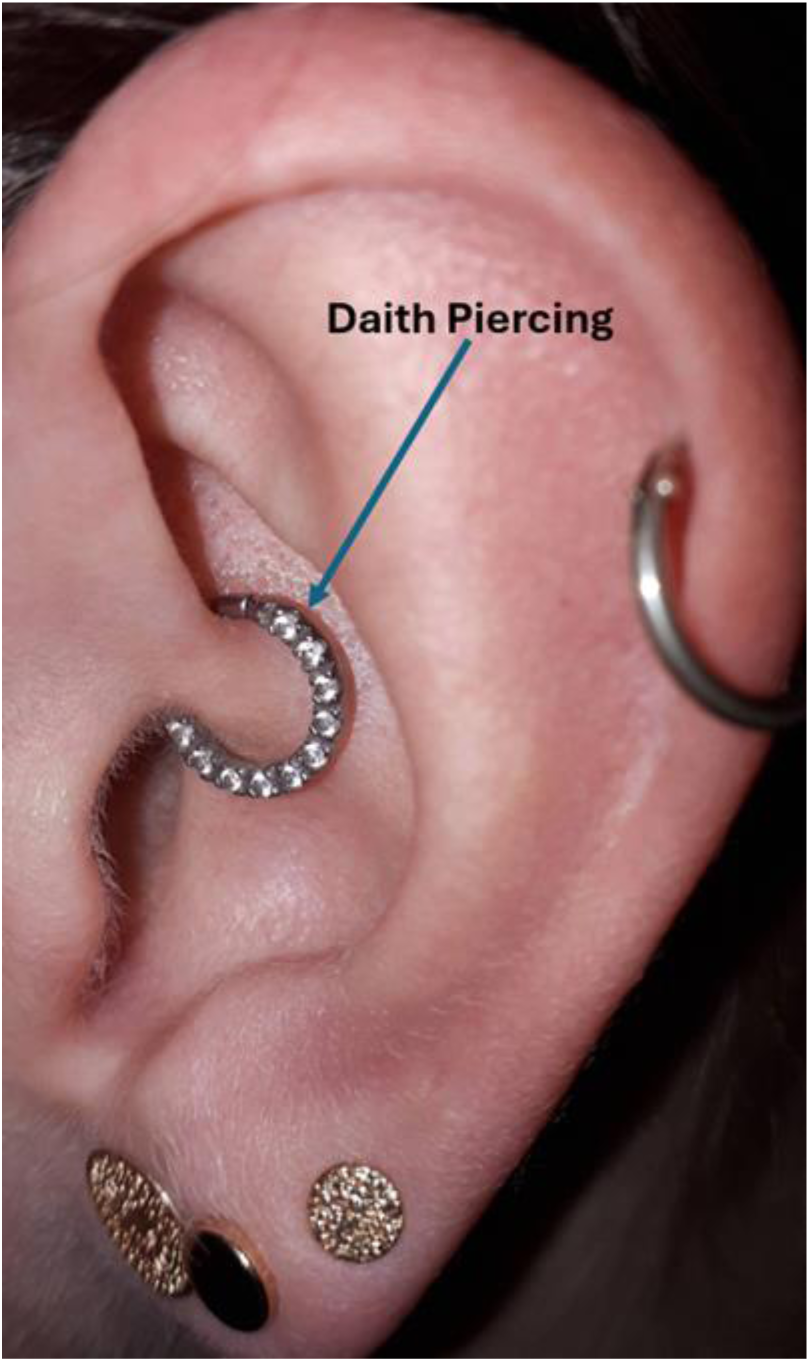
Daith piercing on the crus of the helix.

## Research Question

1) What is the current state of quality and reliability of published YouTube videos concerning the efficacy of DP in relieving migraines?
2) What are the reported characteristics of migraines and the efficacy of DP for migraine treatment? Do the analyzed videos recommend DP for migraine?

## Methods

This systematic video review was conducted in accordance with the Preferred Reporting Items for Systematic Reviews and Meta-Analyses (PRISMA)^6^ guidelines and was pre-registered at PROSPERO (Nr: CRD42024510089). The PRISMA checklist can be found in the supplemental table S1.

### Search strategy

On January 17, 2024, a YouTube search was initiated on www.youtube.com using the keywords “Daith piercing” AND “migraine.” To streamline the search, the default sorting option was set to filter content by “number of views,” which is likely the most frequent choice in YouTube’s sorting algorithm. Other criteria were also considered: relevance, upload date, view count, and rating. All identified YouTube videos from channel inception until the date of data extraction were screened for eligibility.

### Study eligibility criteria

During the video screening, the following exclusion criteria were applied: (1) videos without comments or audio; (2) advertisements; (3) videos not related to migraine and/or DP; and (4) replicated videos, either in part or in whole. Furthermore, videos containing multiple sequences or segments were treated as a single entity, where each video, regardless of its internal divisions, was analyzed as a unified whole rather than as separate parts.

### Data extraction

The retrieval process was facilitated via the YouTube Data application programming interface version 3 (API v3) and programmed on Python, handling queries and storing the results into a pre-defined data file. Two investigators (S.K.P. and G.T. trained in headache rehabilitation) independently evaluated each video by following a pre-specified procedure and standardized form. Initially, the videos underwent full-length screening and were selected based on whether they met the inclusion criteria.

Before the video evaluation and in order to minimize inter-rater variability, both raters scored the first ten videos independently, and any potential issues and questions were discussed.

### Quality assessment

Quality assessment was performed using the Global Quality Scale (GQS).^7^ The GQS involves a 5-point Likert scale to evaluate each video blog’s quality, flow, and user-friendliness. A rating of 1 (very poor) stands for “poor quality, poor flow of the site, most information missing, not at all useful for patients”; while a rating of 2 (poor) means “generally poor quality and poor flow, some information listed but many important topics missing, of very limited use to patients”; 3 (moderate) stands for “moderate quality, suboptimal flow, some important information is adequately discussed but others poorly discussed, somewhat useful for patients”; 4 (good) means “good quality and generally good flow, most of the relevant information is listed”; and 5 (excellent) represents “excellent quality and excellent flow, very useful for patients”.^7^

### Reliability assessment

Reliability assessment was applied by using the DISCERN tool.^8^ DISCERN is structured into three sections consisting of 16 questions aimed at assessing the accuracy of health data for laypersons.^9^ The first eight questions - section one - evaluate a publication’s reliability regarding treatment information; questions 9 to 15 - section two - focus on specific treatment details, and the final question - section three - gives an overall quality rating based on the previous responses.^8^ Each query is assessed on a 5-point scale ranging from 1 (“criterion is not met at all”) to 5 (“criterion is fully met”). A maximum of 75 points could be achieved when, as suggested by Weil et al., we omit the final question and categorize the DISCERN score as followed: 63-75 points for excellent, 51-62 for good, 39-50 for fair, 27-38 for poor, and 16-26 for very poor quality.^10,11^

### Primary and Secondary Outcomes

The pertinent baseline details for each selected video were recorded, including URL, title, channel name, channel handle, channel ID, posted date and time, views, days on YouTube, likes, duration of the video in seconds, and the access date.

The primary outcome of this study was to evaluate the overall quality and reliability of each selected video. This was performed by using two standardized tools. The GQS^7^ was applied to rate the videos on a 5-point scale, where 1 indicates poor quality and 5 reflects excellent quality. Additionally, the DISCERN^8^ tool was used to assess the reliability of the health information provided in the videos, with 16 questions, each rated on a 5-point scale, aimed at evaluating the quality of the health content presented.

As secondary outcomes of interest we defined the efficacy of DP in relieving migraine by rated using a 4-point scale: 1 (very effective), 2 (effective), 3 (less effective), and 4 (not effective at all). Migraine relapse time was also recorded, documenting the time until migraine recurrence after receiving a DP. Healing time was assessed by noting the duration required for the pierced side to heal. Any comparisons made between DP and acupuncture in the videos were also documented. In addition, video updates were recorded, noting whether any updates or edits had been made to the video content after its initial posting.

The number of days the video had been available on YouTube was calculated. Information about which ear(s) were pierced was also recorded, specifying whether the piercing was performed uni- or bilaterally. The frequency of migraines before and after DP was documented, noting any changes in the number of migraine episodes reported by the video participants. Furthermore, the use of rescue medication before and after DP was tracked, as was the use of pharmaceutical pain relief, documenting whether a prescribed medication was utilized to manage migraine pain.

The quality of life (QoL) after DP was assessed on a scale from −2 (much worse) to 2 (much better), reflecting the participant’s perceived improvement or decline in QoL post-procedure. The infection rate was captured, presenting the occurrence or percentage of infections associated with DP. We also noted whether medical consultation for infection was sought and whether the video provider recommended DP as a treatment for migraines. Additionally, any adverse or serious adverse events related to the DP procedure were documented.

Pain intensity measures were included in the analysis whenever possible. Migraine pain intensity was assessed using a Numeric Rating Scale (NRS) ranging from 0 to 10, where 0 represented no pain, and 10 the worst imaginable pain.^12^ Comparisons of pain levels before and after DP were made utilizing this tool. Additionally, pre-existing pain before receiving the procedure was measured on the same NRS scale from 0 to 10.

### Assumptions for Missing or Unclear Data

In instances where specific data points or outcome measures were missing or unclear, several assumptions were made. If a critical piece of data, namely NRS migraine pain pre-DP and post-DP, was not specified in the video, it was marked as “not available.” Similarly, if the video did not provide detailed or full information, e.g., on the use of rescue medication or other relevant details, these were noted as missing but did not affect the overall analysis. Missing data was not replaced.

### Statistical Analysis

To allow detailed insights into the quality and reliability of included videos, these were stratified by the sources: *Health Professionals*, *Personal Experience* or *Other*. The category *Other* included videos from bloggers, piercers and other non-health professionals reporting about DP without personal experience. Due to the non-normal data distribution in the majority of outcomes, descriptive data used to describe the characteristics of the included videos about DP in treating migraine are presented with median (interquartile ranges [IQR]). For binominal outcomes, number and proportions were calculated and compared using Chi-Square statistics or Fisher Exact Test. Continuous outcomes were compared between sources by using Mann-Whitney two-sample statistics. Statistical and graphical analyses were performed using RStudio (R version 2024.04.0), STATA version 15.1 and Sigmaplot version 15. A p-value <0.05 was considered to reflect statistical significance.

### Ethical considerations

Since all obtained videos were publicly available and data were treated confidentially, no personal information was obtained, thus rendering ethical permission unnecessary. Written informed consent to publish photo-documentation was obtained from the patient (Figure 1).

## Results

A total of 496 video recordings were initially identified on YouTube. After removing 20 (4%) duplicate videos, 476 (96%) records were screened, resulting in the exclusion of 230 (48%) videos. Reasons for the exclusion were the lack of comments and audio (n = 102, 21%); videos unrelated to migraines (n = 98, 21%); language restrictions (n = 17, 4%); or videos did not pertain to DP (n = 13, 3%) (Figure 2). Therefore, a total of 246 videos were included in the final analysis (Supplement, eTable 1). Of these, 69 (39.8%) were categorized as *Personal Experience*, 1 (0.5%) as *Healthcare Professionals*, and the remaining 176 as *Other*: The *Other* category contained 63 (25.6%) videos from bloggers, 69 (28.0%) from piercers, and 15 (6.1%) from news channels. The median number (IQR) of days the videos were available on YouTube was 63 (43 ; 78) days, with a median video duration of 3:59 (2:32 ; 8:04) minutes. Table 1 provides an overview of the included video characteristics related to DP for migraine treatment.

**Figure 2:**
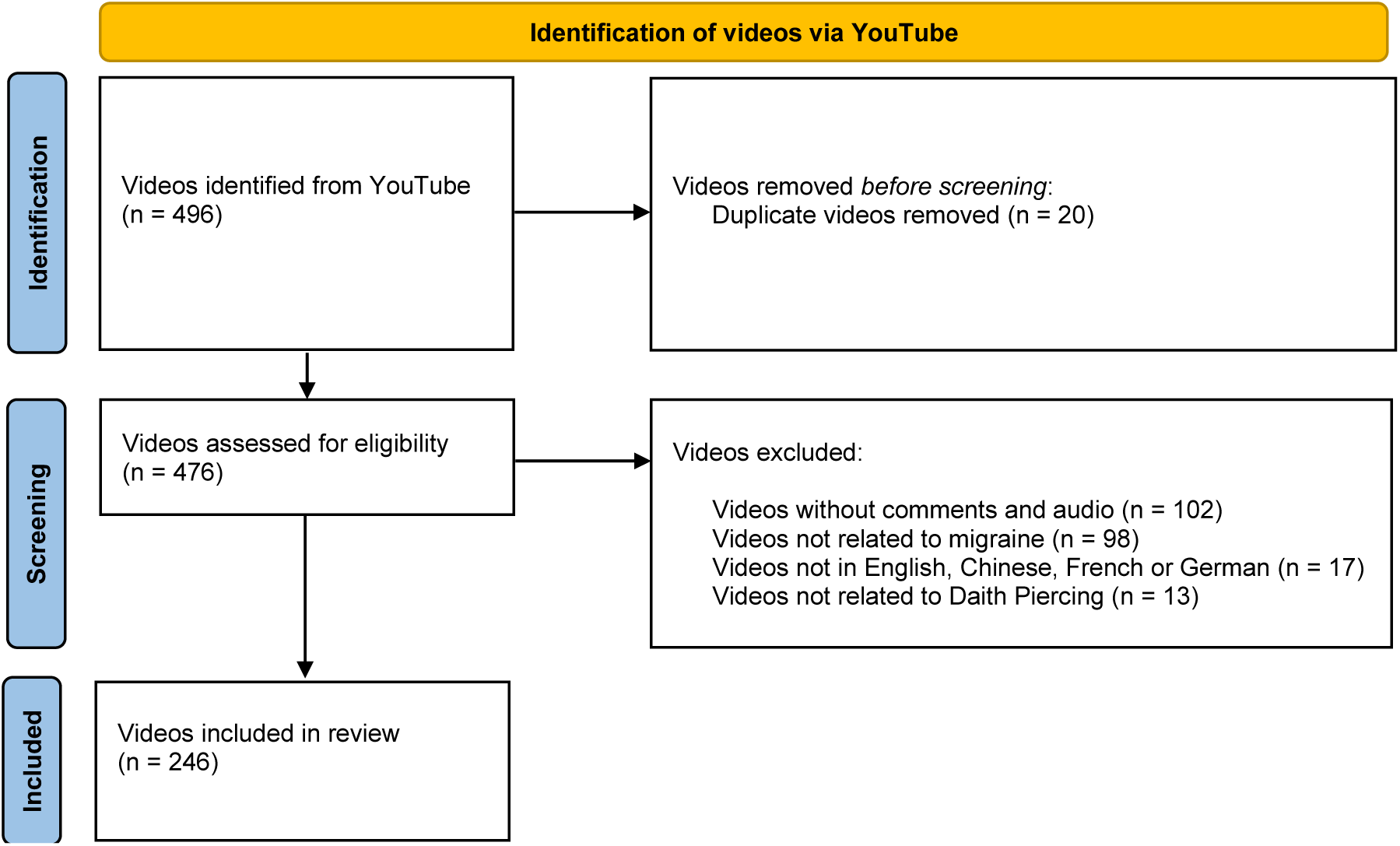
Video selection flowchart.

**Table 1.**
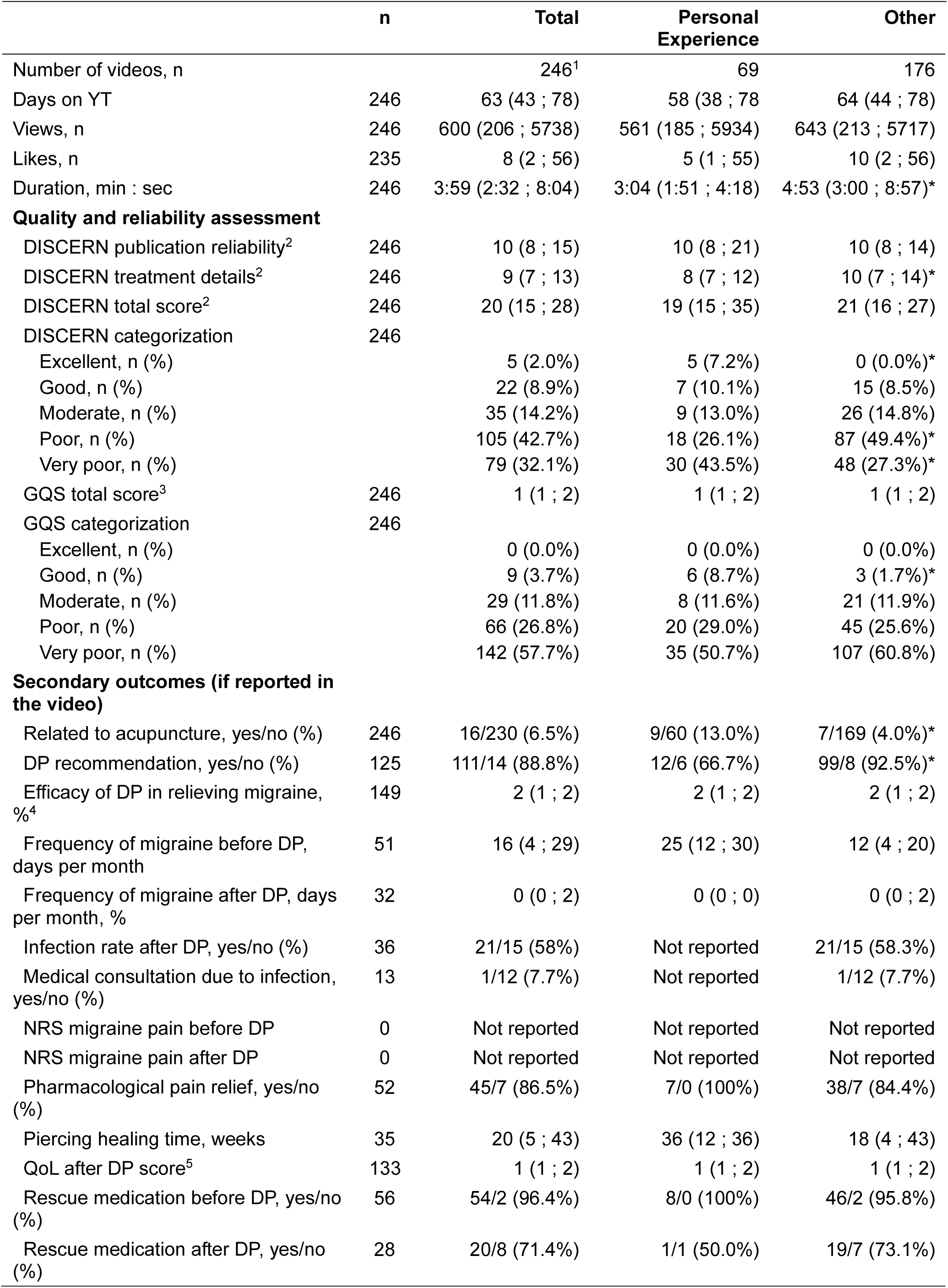
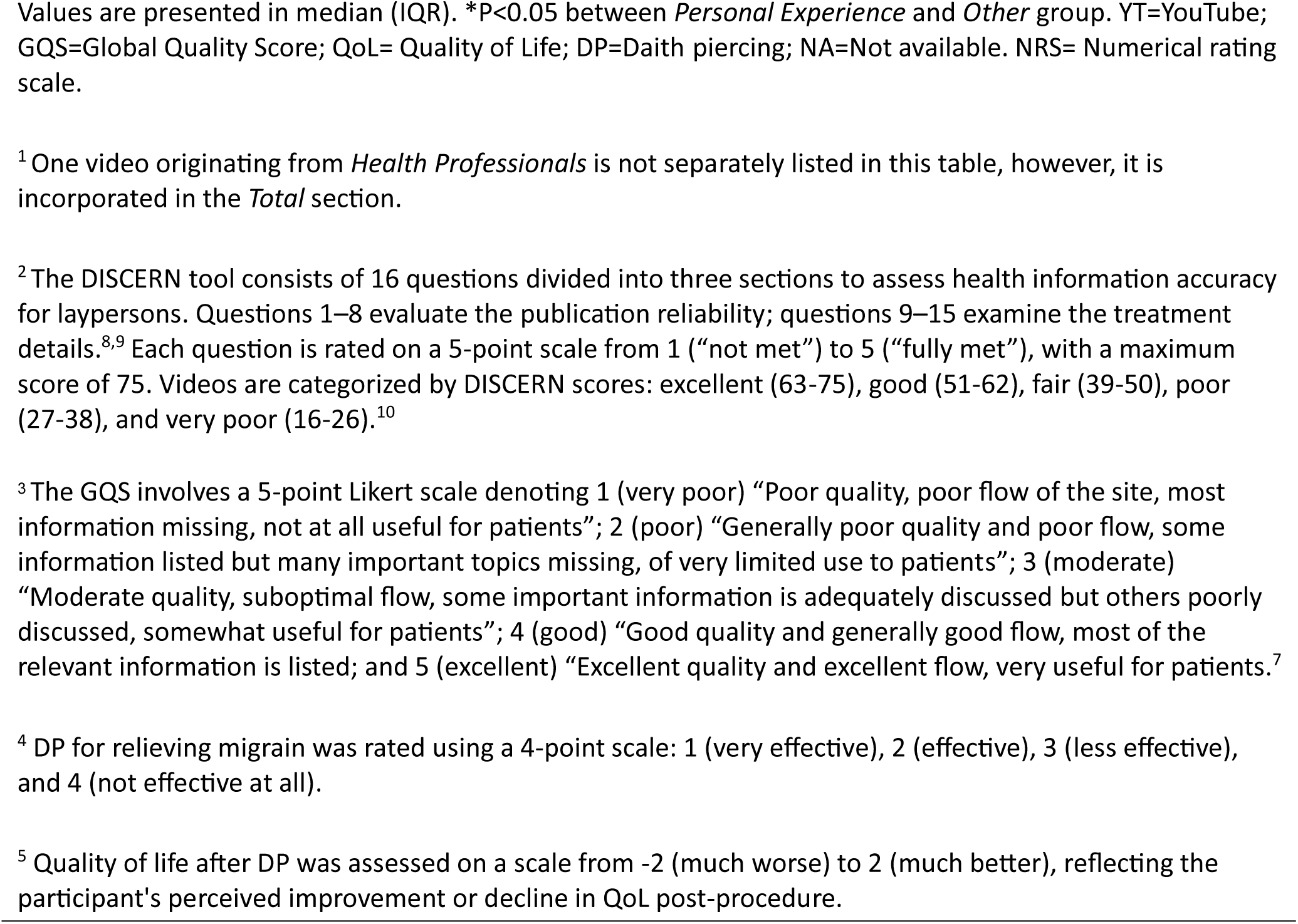
YouTube video characteristics about Daith piercing in treating migraine.

|  | **n** | **Total** | **Personal Experience** | **Other** |
| --- | --- | --- | --- | --- |
| Number of videos, n |  | 246<sup>1</sup> | 69 | 176 |
| Days on YT | 246 | 63 (43 ; 78) | 58 (38 ; 78) | 64 (44 ; 78) |
| Views, n | 246 | 600 (206 ; 5738) | 561 (185 ; 5934) | 643 (213 ; 5717) |
| Likes, n | 235 | 8 (2 ; 56) | 5 (1 ; 55) | 10 (2 ; 56) |
| Duration, min : sec | 246 | 3:59 (2:32 ; 8:04) | 3:04 (1:51 ; 4:18) | 4:53 (3:00 ; 8:57)* |
| **Quality and reliability assessment** |  |  |  |  |
| DISCERN publication reliability<sup>2</sup> | 246 | 10 (8 ; 15) | 10 (8 ; 21) | 10 (8 ; 14) |
| DISCERN treatment details<sup>2</sup> | 246 | 9 (7 ; 13) | 8 (7 ; 12) | 10 (7 ; 14)* |
| DISCERN total score<sup>2</sup> | 246 | 20 (15 ; 28) | 19 (15 ; 35) | 21 (16 ; 27) |
| DISCERN categorization | 246 |  |  |  |
| Excellent, n (%) |  | 5 (2.0%) | 5 (7.2%) | 0 (0.0%)* |
| Good, n (%) |  | 22 (8.9%) | 7 (10.1%) | 15 (8.5%) |
| Moderate, n (%) |  | 35 (14.2%) | 9 (13.0%) | 26 (14.8%) |
| Poor, n (%) |  | 105 (42.7%) | 18 (26.1%) | 87 (49.4%)* |
| Very poor, n (%) |  | 79 (32.1%) | 30 (43.5%) | 48 (27.3%)* |
| GQS total score<sup>3</sup> | 246 | 1 (1 ; 2) | 1 (1 ; 2) | 1 (1 ; 2) |
| GQS categorization | 246 |  |  |  |
| Excellent, n (%) |  | 0 (0.0%) | 0 (0.0%) | 0 (0.0%) |
| Good, n (%) |  | 9 (3.7%) | 6 (8.7%) | 3 (1.7%)* |
| Moderate, n (%) |  | 29 (11.8%) | 8 (11.6%) | 21 (11.9%) |
| Poor, n (%) |  | 66 (26.8%) | 20 (29.0%) | 45 (25.6%) |
| Very poor, n (%) |  | 142 (57.7%) | 35 (50.7%) | 107 (60.8%) |
| **Secondary outcomes (if reported in the video)** |  |  |  |  |
| Related to acupuncture, yes/no (%) | 246 | 16/230 (6.5%) | 9/60 (13.0%) | 7/169 (4.0%)* |
| DP recommendation, yes/no (%) | 125 | 111/14 (88.8%) | 12/6 (66.7%) | 99/8 (92.5%)* |
| Efficacy of DP in relieving migraine, %<sup>4</sup> | 149 | 2 (1 ; 2) | 2 (1 ; 2) | 2 (1 ; 2) |
| Frequency of migraine before DP, days per month | 51 | 16 (4 ; 29) | 25 (12 ; 30) | 12 (4 ; 20) |
| Frequency of migraine after DP, days per month, % | 32 | 0 (0 ; 2) | 0 (0 ; 0) | 0 (0 ; 2) |
| Infection rate after DP, yes/no (%) | 36 | 21/15 (58%) | Not reported | 21/15 (58.3%) |
| Medical consultation due to infection, yes/no (%) | 13 | 1/12 (7.7%) | Not reported | 1/12 (7.7%) |
| NRS migraine pain before DP | 0 | Not reported | Not reported | Not reported |
| NRS migraine pain after DP | 0 | Not reported | Not reported | Not reported |
| Pharmacological pain relief, yes/no (%) | 52 | 45/7 (86.5%) | 7/0 (100%) | 38/7 (84.4%) |
| Piercing healing time, weeks | 35 | 20 (5 ; 43) | 36 (12 ; 36) | 18 (4 ; 43) |
| QoL after DP score<sup>5</sup> | 133 | 1 (1 ; 2) | 1 (1 ; 2) | 1 (1 ; 2) |
| Rescue medication before DP, yes/no (%) | 56 | 54/2 (96.4%) | 8/0 (100%) | 46/2 (95.8%) |
| Rescue medication after DP, yes/no (%) | 28 | 20/8 (71.4%) | 1/1 (50.0%) | 19/7 (73.1%) |

Values are presented in median (IQR). \*P<0.05 between *Personal Experience* and *Other* group. YT=YouTube; GQS=Global Quality Score; QoL= Quality of Life; DP=Daith piercing; NA=Not available. NRS= Numerical rating scale.
<sup>1</sup> One video originating from *Health Professionals* is not separately listed in this table, however, it is incorporated in the *Total* section.
<sup>2</sup> The DISCERN tool consists of 16 questions divided into three sections to assess health information accuracy for laypersons. Questions 1–8 evaluate the publication reliability; questions 9–15 examine the treatment details.<sup>8,9</sup> Each question is rated on a 5-point scale from 1 (“not met”) to 5 (“fully met”), with a maximum score of 75. Videos are categorized by DISCERN scores: excellent (63-75), good (51-62), fair (39-50), poor (27-38), and very poor (16-26).<sup>10</sup>
<sup>3</sup> The GQS involves a 5-point Likert scale denoting 1 (very poor) “Poor quality, poor flow of the site, most information missing, not at all useful for patients”; 2 (poor) “Generally poor quality and poor flow, some information listed but many important topics missing, of very limited use to patients”; 3 (moderate) “Moderate quality, suboptimal flow, some important information is adequately discussed but others poorly discussed, somewhat useful for patients”; 4 (good) “Good quality and generally good flow, most of the relevant information is listed; and 5 (excellent) “Excellent quality and excellent flow, very useful for patients.”<sup>7</sup>
<sup>4</sup> DP for relieving migraine was rated using a 4-point scale: 1 (very effective), 2 (effective), 3 (less effective), and 4 (not effective at all).
<sup>5</sup> Quality of life after DP was assessed on a scale from -2 (much worse) to 2 (much better), reflecting the participant's perceived improvement or decline in QoL post-procedure.

### Primary outcome

#### Quality assessment by GQS

Overall, videos categorized as “very poor” accounted for 57.7% (n = 142) of the total, including 50.7% (n = 35) in the *Personal Experience* group and 60.8% (n = 107) in the *Other* group. “Poor quality” videos made up for 26.8% (n = 66) of all videos, with 29.0% (n = 20) from *Personal Experience* and 25.6% (n = 45) from *Other* as well as the one *Healthcare Professional*s video. No videos were rated as “excellent quality”, and a small proportion of videos were “good quality”, specifically 3.7% (n = 9) overall, 8.7% (n = 6) in *Personal Experience*, and 1.7% (n = 3) in *Other* (P<0.05 compared to the proportion of “good quality” videos in the category *Personal Experience*).

#### Reliability assessment by DISCERN

The analysis of video reliability based on the DISCERN score categorized 42.7% (n = 105) of all videos as having “poor” and 32.1% (n = 79) as “very poor” reliability. In *Personal Experience*, 43.5% (n = 30) were rated “very poor”, while 26.1% (n = 18) scored poor reliability. In *Other*, 27.3% (n = 48) were “very poor”, and 49.4% (n = 87) were “poor quality”. Only 7.2% (n = 5) of *Personal Experience* videos and 8.5% (n = 15) in the *Other* group scored “excellent” reliability.

#### Secondary outcome

The association between DP and acupuncture, rooted in the theoretical framework of traditional acupuncture, was mentioned in 6.5% (n = 16) videos. The left ear was pierced in 26.8% (n = 66) of videos, the right ear in 23.2 % (n = 57), and both ears in 24.8% (n = 61), while 25.2% (n = 62) of videos provided no information as for the pierced side(s) (Table 1). The mean ± SD efficacy score in relieving migraine was 1.8 ± 0.7 points on a 4-point Likert scale, derived from 60.6 % (n = 149) of videos. The average migraine frequency per day/month before DP was 15 ± 11 and decreased to 2 ± 6 after DP. Rescue medication use before DP was described in 21.9% of videos and decreased to 8.1% after DP. The mean duration for piercing healing was 23 ± 18 weeks. The incidence of infection following DP was reported in 8.5% (n = 21) of videos. Only 0.4% (n = 1) of videos described a patient seeking medical consultation for infection, which can be regarded as an adverse event. From all videos providing a recommendation statement (N = 125), 88.8% recommended and 11.2% discouraged DP for migraine relief. According to our statistics, significantly fewer videos in the *Personal Experience* recommend DP for migraines compared to those in the *Other* category.

## Discussion

An increasing number of migraine patients are turning to social media, such as YouTube, as a source of information regarding DP in migraine treatment, which is largely attributable to the platform’s ease of access. The 246 videos analyzed in this systematic video review collectively received a total of approximately 4.7 million views, indicating a substantial interest in DP as a possible therapeutic intervention for migraine. Overall, 88.8% of videos recommended DP to relieve migraine. In contrast and based on our systematic video review, YouTube cannot be recommended from a medical point of view for patients seeking information on DP as a treatment for migraine. The poor quality and reliability of the video content, high rates of missing data and limited involvement of healthcare professionals raise concerns about the information quality, the accuracy of the content, and reliability of the information available on YouTube regarding DP for migraine treatment.

In the study by Karacan et al. on Botox treatment, crucial correlations were observed between GQS and DISCERN (R = 0.757; P <0.001), with an average DISCERN score of 3.09 out of 5.^13^ The present DP study showcased “poor” and “very poor” quality DISCERN scores across most video categories, with 42.7% of total videos rated as “poor” and 32.1% as “very poor”. Specifically, 27.3% of videos from *Other* and 43.5% from *Personal Experience* were ranked as “very poor” quality, with only 7.2% from *Personal Experience* being evaluated as “excellent”. The sole video from a *Healthcare Professionals* in the DP analysis was also graded as “very poor”. As for reliability, the videos from Karacan er al.,showed moderate DISCERN scores, with substantial differences observed based on the narrator’s qualifications (P = 0.002), whereas videos produced by university/nonprofit physicians or professional organizations pointed out greater reliability.^13^ While videos on Botox for migraines displayed higher quality and reliability,^13^ DP videos largely lacked reliable content.

In another study, Hakyemez et al. demonstrated that videos uploaded by physicians had a notable higher GQS (4.66 ± 0.47) compared to those uploaded by non-physicians (3.30 ± 0.91, P <0.01). Similarly, DISCERN scores were higher for physician-uploaded videos (4.56 ± 0.50) compared to non-physician videos (3.32 ± 0.85, P < 0.01).^14^ In comparison, this DP study discovered a high prevalence of poor quality videos and an overall mean GQS score of (1.61 ± 0.83) and DISCERN score of 24 ± 10 in the 246 included videos. While both studies evaluated the quality and reliability of YouTube videos, substantial differences were observed. Neonatal sepsis videos, particularly those submitted by healthcare professionals, confirmed notably higher GQS and DISCERN scores.^14^ Again, DP videos exhibited much lower scores in GQS and DISCERN scores.

The majority of the videos analyzed by Erten et al. on glycated hemoglobin levels (HbA1c) involved professionals, with 34% originating from medical educational channels, 25% from medical doctors, and 25% from other healthcare providers.^15^ The GQS for all videos was 3.7 ± 1.0 (median: 4), with medical doctor-uploaded videos achieving the highest GQS scores (4.6 ± 0.4, median: 5).^15^ The mean DISCERN score across all videos was 59.0 ± 10.5 (median: 60.5), with videos produced by medical doctors again scoring the highest scores (66 ± 8, median: 68). A considerable correlation was observed between GQS and DISCERN scores (R = 0.874; P <0.05).^15^ Furthermore, regression analysis revealed that the video duration was profoundly associated with GQS and DISCERN scores (P <0.05).^15^ In contrast, the current DP study predominantly included videos from non-professional sources, highlighting the potential for improvement.

The findings of this study suggest that YouTube does not provide high quality and reliable information regarding DP as a treatment for migraine. The qualitative content and the reliability of information on YouTube regarding DP for migraine are poor across all video categories, as displayed by both GQS and DISCERN scores (Figure 3). Out of the 246 videos analyzed, the vast majority (n = 245) were created based on personal experience or by bloggers, piercers or news channels without providing sufficient medical information. This high prevalence of non-medical content may be explained by the subjective nature of migraine pain, which patients primarily experience themselves. Consequently, videos are more likely produced by individuals sharing personal narratives and experiences rather than by medical professionals. Although higher quality ratings of videos originating from *Personal Experience* were observed, the overall poor quality and trend raises concerns about the potential for misinformation related to DP for migraine treatment on YouTube.

**Figure 3:**
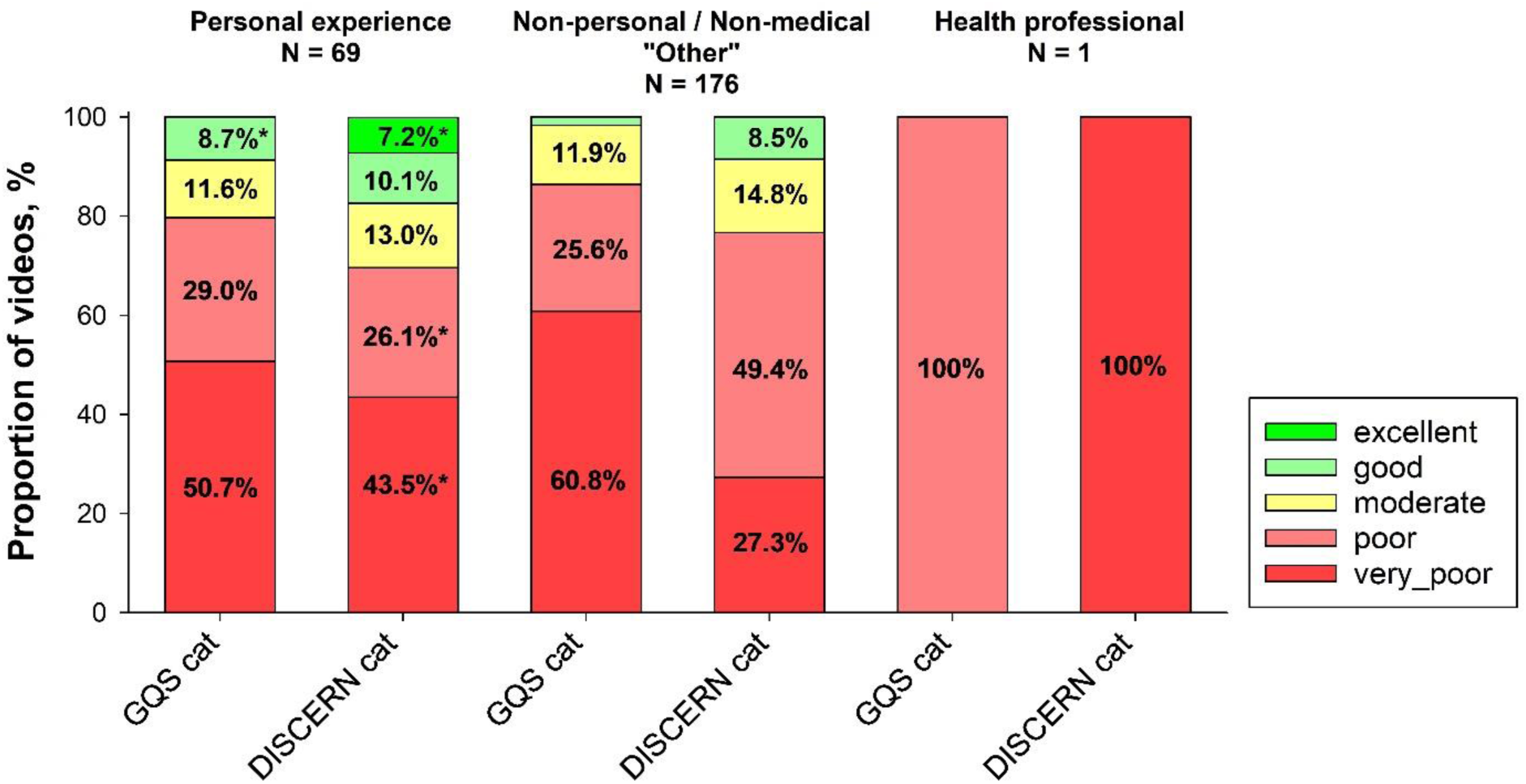
Quality and reliability assessments by the Global Quality Scale (GQS) and DISCERN categorization across the three video categories: *Personal experience*, *Other*, and *Health professionals*. The category *Other* contains videos published from bloggers, piercers and news channels. *P<0.05 versus *Personal Experience* in the same quality category of the GQS or DISCERN score.

Despite some videos reporting migraine relief from DP, the majority of these miss out on reliable information, suggesting a publication bias. The reported reduction in migraine frequency after DP - from an average of 20.7 days per month to 13.0 attacks/month - might underscore the potential of DP as an alternative treatment option in migraine management. However, the high rates of missing data and the lack of prospective clinical trials do not allow to recommend DP for migraine. Furthermore, Pradhan et. al. have demonstrated in their literature review that no link between auricular acupuncture and DP exists in the context of migraine relief,^4^ raising further concerns about the efficacy and mechanism of action of DP for migraine relief.

### Limitation

The current study has some limitations. First, numerous other social media platforms such as Facebook, TikTok, Instagram, LinkedIn, Twitter, Vimeo, and other video platforms provide similar content. Therefore, the inclusion of additional platforms might have provided different results. Nevertheless, the scope of this study focused on YouTube, since YouTube supports videos with a longer duration, potentially allowing the creators to discuss the topic in full. Nonetheless, future research could consider examining other social media platforms to better understand how social media serves as a source of information about DP in the context of migraine management.^16^ Furthermore, the outcome of this study may be influenced by the search terms selected. Our systematic video analysis focused on the keywords “Daith piercing” AND “Migraine.” However, these may not necessarily reflect the keywords that an average user might enter while exploring content on YouTube on this relevant subject. It is possible that patients could utilize different terms, leading to different video results and findings. Lastly, analyzing video content is a subjective interpretation, which might result in varying quality assessments.

## Conclusion

Based on our systematic video review, YouTube cannot be recommended from a medical point of view for patients seeking information on DP as a treatment for migraine. The poor quality and reliability of the video content, high rates of missing data, insufficient updates, and limited involvement of healthcare professionals raise concerns about the information quality, the accuracy of the content, and reliability of the information available on YouTube regarding DP for migraine treatment. Furthermore, most videos reflect anecdotal experiences rather than scientifically validated data, which might lead to a positive reporting bias. A large number of videos focus on immediate headache relief experience, often neglecting the long-term consequences of DP healing time, infections and potential side effects.

## Supporting information

Suppement

## Data Availability

All data produced in the present study are available upon reasonable request to the authors

## Acknowledgment

This review was supported by SWISS TCM UNI and TCM Ming Dao AG. The supporting sources had no role in designing this study, in writing the manuscript, or in deciding to submit this manuscript.

## Author Contributions

**Saroj K. Pradhan**: Conceptualization; data curation; formal analysis; investigation; methodology; project administration; validation; writing – original draft; writing – review and editing. **Michael Furian**: Conceptualization; data curation; formal analysis; investigation; methodology; supervision; validation; writing – review and editing. **Giada Todeschini**: data curation; Formal analysis; investigation; writing – review and editing. **Qiong Schürer**: data curation; Formal analysis; investigation; writing – review and editing. **Xiaoying Wang**: Formal analysis; investigation; writing – review and editing. **Bingjun Chen**: Formal analysis; investigation; writing – review and editing. **Yiming Li**: Formal analysis; investigation; writing – review and editing. **Andreas R. Gantenbein**: Conceptualization; formal analysis; investigation; methodology; supervision; validation; writing – review and editing.

All authors have read and agreed to the published version of the manuscript.

## Abbreviations

DP: Daith piercing
IQR: interquartile ranges
GQS: Global Quality Score
QoL: quality of life
NA: Not available
NRS: Numerical rating scale

## Notes

**Conflict of Interest Statement:** No conflict

### Competing Interest Statement

The authors have declared no competing interest.

### Funding Statement

This review received no external funding.

