## Supplementary material for "Daith piercing, a social media hype on YouTube for the treatment of migraine? A systematic video analysis of quality and reliability": Suppement

**Conflict of Interest Statement:** No conflict

**Keywords:** Pain, headache disorders, migraine, piercing, YouTube, complementary and alternative medicine,

**Funding:** This review received no external funding.

| ***eTable 1*** | |  |  |  |  |  |  |  |  |
| --- | --- | --- | --- | --- | --- | --- | --- | --- | --- |
| ***Index*** | ***URL*** | **source** | **Title** | **Upload date** | **Views** | **Likes** | **Duration, sec** | **DISCERN_cat** | **GQS_cat** |
| *1* | *https://www.youtube.com/watch?v=2yl1KhmxqSI* | Health Professional | Tracy Perkins - Vagus Nerve Stimulation Therapy via Daith Piercing | 09.02.2021 | 498 | 8 | 96 | Very poor | Poor |
| *2* | *https://www.youtube.com/watch?v=NnHETlsvYHA* | Personal experience | DID THE DAITH PIERCING HELP MY MIGRAINES? | 04.02.2020 | 8324 | 184 | 424 | Good | Good |
| *3* | *https://www.youtube.com/watch?v=jblWb0Put8Y* | Personal experience | Do Daith Piercings help with Migraine? A long term review. 3 Years Later. | 02.06.2020 | 6107 | 69 | 611 | Good | Good |
| *4* | *https://www.youtube.com/watch?v=8ay-9wL8ERo* | Personal experience | The Whole Truth - Daith Piercing | 02.07.2020 | 119634 | 3468 | 279 | Good | Moderate |
| *5* | *https://www.youtube.com/watch?v=pWEzeYvdVqM* | Personal experience | Comprehensive Guide to Daith Piercings | 09.03.2021 | 50123 | 1377 | 898 | Good | Moderate |
| *6* | *https://www.youtube.com/watch?v=k4Cari1IntI* | Personal experience | This Piercing Claims To Get Rid of Migraines &amp; Headaches!? *IS IT TRUE?* | 25.03.2020 | 10389 |  | 344 | Good | Moderate |
| *7* | *https://www.youtube.com/watch?v=dCCXqmwAsLk* | Personal experience | An ear piercing...curing migraines? Some people say theirs are doing just that | 18.05.2019 | 2772 | 9 | 167 | Good | Moderate |
| *8* | *https://www.youtube.com/watch?v=zql_6tFqImE* | Personal experience | Daith Piercings &amp; Migraines\| My Opinion \|NativeBeauty | 12.04.2019 | 1741 | 31 | 317 | Good | Moderate |
| *9* | *https://www.youtube.com/watch?v=5dsnTZHCEgs* | Personal experience | DID THE DAITH PIERCING HELP MY MIGRAINES?? \| Migraine Update 10 Months with the Daith Ear Piercing | 10.06.2020 | 1280 | 30 | 509 | Good | Moderate |
| *10* | *https://www.youtube.com/watch?v=X7_Lq0DP7IY* | Personal experience | Daith Piercing Update + Many Natural Migraine Relief Ideas! | 07.11.2018 | 792 | 21 | 714 | Good | Moderate |
| *11* | *https://www.youtube.com/watch?v=la33Pcr2FBI* | Personal experience | Using a piercing to get rid of migraine pain | 13.07.2017 | 2600 | 30 | 178 | Good | Poor |
| *12* | *https://www.youtube.com/watch?v=PtilLcLhtZo* | Personal experience | Piercing away painful migraines | 26.02.2016 | 2051 | 12 | 191 | Good | Poor |
| *13* | *https://www.youtube.com/watch?v=z13B-QbUZZg* | Personal experience | Daith Piercing for Migraines: One Year Update | 27.04.2017 | 1717 | 21 | 279 | Good | Poor |
| *14* | *https://www.youtube.com/watch?v=rN0ppjvVvDE* | Personal experience | PIERCING DAITH LA SOLUTION ANTI MIGRAINE | 08.01.2020 | 1243 | 11 | 580 | Good | Poor |
| *15* | *https://www.youtube.com/watch?v=gV0EAvKbjm8* | Personal experience | Daith piercing for migraines. Did it work? My experience and results! | 08.01.2022 | 657 | 41 | 451 | Good | Poor |
| *16* | *https://www.youtube.com/watch?v=vDe2QMcKWHI* | Personal experience | Daith piercing for migraines | 28.01.2017 | 298 | 4 | 222 | Good | Poor |
| *17* | *https://www.youtube.com/watch?v=CiZuNBbYIwI* | Personal experience | EIN PIERCING GEGEN MIGRÄNE ?!? \| MIGRÄNEPIERCING \| Erfahrungsbericht nach 1 Jahr \| Daithpiercing | 24.11.2017 | 33446 | 355 | 1509 | Fair | Good |
| *18* | *https://www.youtube.com/watch?v=gTZtH4BbfqU* | Personal experience | My Daith Piercing Experience - 1 Month Update \| Does this thing work for migraines..? | 14.02.2017 | 8663 | 58 | 254 | Fair | Moderate |
| *19* | *https://www.youtube.com/watch?v=o8zpAfJY4pI* | Personal experience | Daith piercing and migraines: part 9 | 28.02.2016 | 4098 | 37 | 271 | Fair | Moderate |
| *20* | *https://www.youtube.com/watch?v=VKtr2Nm9-CQ* | Personal experience | My Migraine Daith Piercing Experience \| My FIRST Piercing! + 1 Week Migraine Update | 07.10.2019 | 2897 | 55 | 761 | Fair | Moderate |
| *21* | *https://www.youtube.com/watch?v=n6BbZ0xHPFY* | Personal experience | Daith piercing and migraines: part 11 | 08.05.2016 | 1306 | 15 | 191 | Fair | Moderate |
| *22* | *https://www.youtube.com/watch?v=rLsu0WZ-vFM* | Personal experience | Daith Piercings Cure Migraines?!?!? \| Piercing Reaction \| | 13.10.2021 | 1148 | 23 | 2582 | Fair | Moderate |
| *23* | *https://www.youtube.com/watch?v=VLg9NYhj-t4* | Personal experience | Piercings bring relief for some who suffer from migraine headaches | 24.05.2017 | 42438 | 446 | 241 | Fair | Poor |
| *24* | *https://www.youtube.com/watch?v=Aiv7XTnPXto* | Personal experience | Daith Piercing for Migraines \| 2 1/2 Year Update! | 16.12.2019 | 5695 | 132 | 1349 | Fair | Poor |
| *25* | *https://www.youtube.com/watch?v=3u4luBk19oI* | Personal experience | Daith Piercings and Migraines - 3 Month Update | 12.04.2017 | 1979 | 25 | 298 | Fair | Poor |
| *26* | *https://www.youtube.com/watch?v=ghB6LhPmXKY* | Personal experience | Daith piercing et migraines | 20.07.2021 | 1049 | 6 | 208 | Fair | Poor |
| *27* | *https://www.youtube.com/watch?v=Fbl8IErz0e0* | Personal experience | London Migraine Clinic - Leah&#39;s Medi-Daith for Migraine Journey | 12.09.2022 | 629 | 1 | 385 | Fair | Poor |
| *28* | *https://www.youtube.com/watch?v=J49ziKYY9C0* | Personal experience | DAITH PIERCING UPDATE! Did it help me with my MIGRAINES?? | 23.01.2021 | 597 | 20 | 474 | Fair | Poor |
| *29* | *https://www.youtube.com/watch?v=GGTo1Mejiyo* | Personal experience | Daith Piercing Experience: Migraine Relief + 10 Month Update! - Challenge Accepted | 19.02.2018 | 306 | 5 | 426 | Fair | Poor |
| *30* | *https://www.youtube.com/watch?v=jKFVzFD7M9M* | Personal experience | Daith Piercing For Migraine II SHOCKING Results !!! | 09.11.2017 | 304 | 2 | 486 | Fair | Poor |
| *31* | *https://www.youtube.com/watch?v=ztOaPjucZto* | Personal experience | Daith Piercing \| My reasoning why, Pain scale &amp; first 5 days of healing \| Gabby Thomson (Vlog #203 ) | 10.07.2022 | 253 | 7 | 2565 | Fair | Poor |
| *32* | *https://www.youtube.com/watch?v=R9_LK0lRu_0* | Personal experience | DAITH PIERCING CURE MIGRAINES/MY EXPERIENCE/CHIT CHAT/GRWM | 01.01.2020 | 96 | 9 | 1640 | Fair | Poor |
| *33* | *https://www.youtube.com/watch?v=g-VLfzzXXzc* | Personal experience | Daith Piercings For Migraine with Gary Harrison &amp; Richard Soper | 02.01.2024 | 90 | 1 | 237 | Fair | Poor |
| *34* | *https://www.youtube.com/watch?v=WLE1dPIBMDg* | Personal experience | Do Daith Piercings Help with Migraines? | 27.01.2018 | 77 | 1 | 2191 | Fair | Poor |
| *35* | *https://www.youtube.com/watch?v=FxIknkhfrxw* | Personal experience | DAITH PIERCINGS and migraines - do they work? | 09.05.2017 | 44 | 1 | 553 | Fair | Poor |
| *36* | *https://www.youtube.com/watch?v=ziZ6bpNLvl8* | Personal experience | daith piercing review! (price, pain, &amp; does it help with migraines?) | 17.04.2019 | 39941 | 878 | 386 | Fair | Very poor |
| *37* | *https://www.youtube.com/watch?v=LiyNUbAs8hw* | Personal experience | Daith Piercing for Migraine - 1 Month Update | 13.01.2017 | 11205 | 163 | 536 | Fair | Very poor |
| *38* | *https://www.youtube.com/watch?v=RUslsBeXVVQ* | Personal experience | 3 week daith piercing update | 31.01.2016 | 1931 | 15 | 180 | Fair | Very poor |
| *39* | *https://www.youtube.com/watch?v=nTXAA5cxnDY* | Personal experience | Daith Piercing &amp; Migraines!?!?! | 07.07.2016 | 992 | 4 | 231 | Fair | Very poor |
| *40* | *https://www.youtube.com/watch?v=MJ_t0PZ8sY0* | Personal experience | Daith Piercing for Migraine Sussex - Lindsey&#39;s Reaction | 17.11.2017 | 165 | 1 | 325 | Fair | Very poor |
| *41* | *https://www.youtube.com/watch?v=6Jiap9MDAiY* | Personal experience | 6 Month Daith Piercing For Migraines Update | 30.11.2018 | 76 | 2 | 653 | Fair | Very poor |
| *42* | *https://www.youtube.com/watch?v=d7A3ov1QpEk* | Personal experience | #daithpiercing #migraines | 24.01.2022 | 5 | 1 | 79 | Fair | Very poor |
| *43* | *https://www.youtube.com/watch?v=qYbv-yYR2n8* | Personal experience | My Daith Piercing Story \| Does It Really Work For Migraines?!? | 13.09.2017 | 167370 | 1766 | 2358 | Poor | Moderate |
| *44* | *https://www.youtube.com/watch?v=wmc8NImy4rM* | Personal experience | My Daith Piercing Experience: Migraine Relief (2.16) | 17.03.2016 | 80676 | 549 | 901 | Poor | Moderate |
| *45* | *https://www.youtube.com/watch?v=TlmYmg4K5U8* | Personal experience | One Year Daith Piercing Update For Migraines- Ashley Witmer | 29.04.2018 | 70558 | 847 | 437 | Poor | Moderate |
| *46* | *https://www.youtube.com/watch?v=ynsSVLKqYcY* | Personal experience | Daith Piercing Update + How I Clean My Piercing | 27.08.2018 | 36758 | 545 | 317 | Poor | Moderate |
| *47* | *https://www.youtube.com/watch?v=5djHXpK5Oxc* | Personal experience | MIGRAINE DAITH PIERCING + TRAGUS PIERCING // UPDATE + PIERCING CHANGE | 19.07.2019 | 23023 | 470 | 643 | Poor | Moderate |
| *48* | *https://www.youtube.com/watch?v=54poce3s9Uo* | Personal experience | WILL THIS HURT? Daith Piercing for Migraine Relief | 14.05.2021 | 4138 | 55 | 930 | Poor | Moderate |
| *49* | *https://www.youtube.com/watch?v=9hmgZ57RvoE* | Personal experience | Daith piercing and migraines: part 6 | 23.12.2015 | 4053 | 58 | 144 | Poor | Moderate |
| *50* | *https://www.youtube.com/watch?v=wL8Ei31FJTU* | Personal experience | Do Daith Piercings Fix Migraines? ~ My Experience | 01.05.2017 | 2999 | 52 | 664 | Poor | Moderate |
| *51* | *https://www.youtube.com/watch?v=g4Mn9XK4Luw* | Personal experience | DAITH Piercing for Migraines \| Did it actually work? | 26.02.2022 | 225 | 9 | 1290 | Poor | Moderate |
| *52* | *https://www.youtube.com/watch?v=kHYLA6QATo0* | Personal experience | How to Change Daith Piercing Jewelry | 09.09.2016 | 504063 | 2593 | 275 | Poor | Poor |
| *53* | *https://www.youtube.com/watch?v=YLwEqEEl3KA* | Personal experience | We Get Piercings To Try And Cure Our Chronic Migraines | 18.12.2020 | 380703 | 11105 | 579 | Poor | Poor |
| *54* | *https://www.youtube.com/watch?v=zlGxC0bzHHc* | Personal experience | Getting My 20th Piercing, A Daith Piercing \| Macro Beauty \| Refinery29 | 20.10.2021 | 338081 | 6437 | 382 | Poor | Poor |
| *55* | *https://www.youtube.com/watch?v=6QYijNg52uA* | Personal experience | I GOT MY DAITH PIERCED TO STOP MIGRAINES \| MY EXPERIENCE | 09.06.2018 | 81954 | 3048 | 538 | Poor | Poor |
| *56* | *https://www.youtube.com/watch?v=nuy-08XcJys* | Personal experience | Daith Piercing for Migraine Sussex - Miffy - 2 Months On | 09.02.2018 | 50468 | 15 | 183 | Poor | Poor |
| *57* | *https://www.youtube.com/watch?v=cpv05uL7djo* | Personal experience | Did This Piercing Cure My Migraines? -Daith Piercing Review!- | 15.11.2016 | 49649 | 1257 | 245 | Poor | Poor |
| *58* | *https://www.youtube.com/watch?v=OphJnM_d9f0* | Personal experience | Daith Piercing for Migraine Sussex - Holly Reaction - Re-Stimulation of Vagus Nerve | 06.03.2018 | 34845 | 11 | 310 | Poor | Poor |
| *59* | *https://www.youtube.com/watch?v=2EnMsQs14DE* | Personal experience | PROS &amp; CONS: Daith Piercing \|NativeBeauty | 18.10.2018 | 27771 | 263 | 573 | Poor | Poor |
| *60* | *https://www.youtube.com/watch?v=BNk0ZM9345w* | Personal experience | DAITH PIERCING MIGRANE - PAIN?,COST?,HEALING AND MARIA TASH EXPERIENCE | 05.12.2021 | 14270 | 346 | 1025 | Poor | Poor |
| *61* | *https://www.youtube.com/watch?v=VkvwDRg_yXc* | Personal experience | Curing Migraines With a DAITH Piercing | 18.02.2020 | 3345 | 174 | 515 | Poor | Poor |
| *62* | *https://www.youtube.com/watch?v=tIhw9fJCyHI* | Personal experience | Daith Piercing for Migraine Sussex - Lindsey - 2 Months After Treatment | 09.01.2018 | 524 | 4 | 212 | Poor | Poor |
| *63* | *https://www.youtube.com/watch?v=pone1iaZPvY* | Personal experience | Migraine Piercing for the lovely Mariane from Brazil | 30.11.2020 | 499 | 16 | 224 | Poor | Poor |
| *64* | *https://www.youtube.com/watch?v=ROKFa3IOfks* | Personal experience | Daith Piercing for Migraine Sussex - Nigel - 4 Months After Treatment | 09.01.2018 | 496 | 4 | 211 | Poor | Poor |
| *65* | *https://www.youtube.com/watch?v=CK1DLPQJRFY* | Personal experience | Daith Piercing for Migraine Sussex - Jonathan D Ellis - Actor &amp; Musician | 12.01.2018 | 394 | 3 | 227 | Poor | Poor |
| *66* | *https://www.youtube.com/watch?v=_monP8cViqs* | Personal experience | Tears for Anita at Daith Medical Ltd, 22 Harley Street today. She tells her Daith Piercing story | 07.02.2022 | 387 | 2 | 88 | Poor | Poor |
| *67* | *https://www.youtube.com/watch?v=3WkKkVIOle4* | Personal experience | Daith Piercing for Migraine Sussex - Jo - Cosmetic Vs Medical Daith Piercing | 09.01.2018 | 384 | 3 | 186 | Poor | Poor |
| *68* | *https://www.youtube.com/watch?v=bWpyezN8fdc* | Personal experience | Daith Piercing cures 10 years of pain for 13 year old Lilly | 17.02.2020 | 298 | 3 | 183 | Poor | Poor |
| *69* | *https://www.youtube.com/watch?v=vHgjzpv3qxo* | Personal experience | month daith piercing/ migraine update | 19.03.2017 | 231 | 3 | 358 | Poor | Poor |
| *70* | *https://www.youtube.com/watch?v=XyaoFv21ZNU* | Personal experience | I GOT MY DAITH PIERCED TO STOP MY MIGRAINES! VLOG | 17.09.2019 | 170 | 10 | 1038 | Poor | Poor |
| *71* | *https://www.youtube.com/watch?v=2p3vgbX7hzg* | Personal experience | Daith piercing update 1 month | 21.05.2016 | 150 | 6 | 119 | Poor | Poor |
| *72* | *https://www.youtube.com/watch?v=Cp1WxEe2Tuk* | Personal experience | Has The Daith Piercing Helped My Migraines? | 13.08.2019 | 106 | 4 | 610 | Poor | Poor |
| *73* | *https://www.youtube.com/watch?v=hc6xkYxTU48* | Personal experience | Daith piercing and trash bags | 11.02.2020 | 78 | 1 | 306 | Poor | Poor |
| *74* | *https://www.youtube.com/watch?v=ZrA7eqDkAGQ* | Personal experience | HEALTH WATCH: Daith Piercing Gaining In Popularity For Migraine Sufferers | 12.01.2019 | 44 | 3 | 221 | Poor | Poor |
| *75* | *https://www.youtube.com/watch?v=Blq-kI-RQJQ* | Personal experience | Migraine Daith Piercing Review #shorts | 30.09.2021 | 31 | 0 | 77 | Poor | Poor |
| *76* | *https://www.youtube.com/watch?v=a4nqC6IW-Eg* | Personal experience | I Get A DAITH PIERCING To Prevent &amp; Stop Migraines Vlog | 23.06.2018 | 122198 | 158 | 689 | Poor | Very poor |
| *77* | *https://www.youtube.com/watch?v=iW8hILmBRg4* | Personal experience | Migraine \| Daith piercing- I fucked up | 12.12.2016 | 115768 | 582 | 1317 | Poor | Very poor |
| *78* | *https://www.youtube.com/watch?v=dHsSGd3MoZY* | Personal experience | WARNING! GRAPHIC CONTENT // The Migraine Daith Piercing \| Vlog | 28.11.2018 | 82251 | 801 | 610 | Poor | Very poor |
| *79* | *https://www.youtube.com/watch?v=ZMNUDMUwXrU* | Personal experience | Caroline&#39;s Migraine Reaction | 13.03.2018 | 80504 | 15 | 256 | Poor | Very poor |
| *80* | *https://www.youtube.com/watch?v=dLtzuXmfmME* | Personal experience | Daith Piercing for Migraine Sussex - Molly - Age 16 Migraines for 10 years | 28.02.2018 | 36540 | 5 | 198 | Poor | Very poor |
| *81* | *https://www.youtube.com/watch?v=snjhMN3-wKY* | Personal experience | Getting a daith piercing for migraines | 09.09.2018 | 29335 | 2445 | 753 | Poor | Very poor |
| *82* | *https://www.youtube.com/watch?v=EyuCtHI4UMQ* | Personal experience | My Daith Piercing Experience: Migraine Relief Update | 19.04.2016 | 26762 | 372 | 442 | Poor | Very poor |
| *83* | *https://www.youtube.com/watch?v=KNL0qLqTDqU* | Personal experience | Daith Piercing - One Month Update\|Migraines, Swelling, Pain \|NativeBeauty | 05.04.2017 | 25791 | 164 | 522 | Poor | Very poor |
| *84* | *https://www.youtube.com/watch?v=xPWBIh77v2E* | Personal experience | Daith piercing and migraines | 16.11.2015 | 24875 | 190 | 351 | Poor | Very poor |
| *85* | *https://www.youtube.com/watch?v=B9O1wDMj5RE* | Personal experience | Does a DAITH Piercing Really Heal Migraines? \| 6 Month Update | 24.04.2018 | 20136 | 287 | 227 | Poor | Very poor |
| *86* | *https://www.youtube.com/watch?v=ac6qZu02zlg* | Personal experience | My DAITH PIERCING Experience \| Natural Headache Pain Reliever | 24.10.2017 | 12102 | 175 | 259 | Poor | Very poor |
| *87* | *https://www.youtube.com/watch?v=RtLkz8Rp9u8* | Personal experience | Daith Piercing for Migraine - 3 Month Update | 10.03.2017 | 10397 | 61 | 518 | Poor | Very poor |
| *88* | *https://www.youtube.com/watch?v=osmFQae33Gw* | Personal experience | Daith Piercing For Migraines Which Side Should I Get It On? | 15.08.2017 | 6307 | 225 | 143 | Poor | Very poor |
| *89* | *https://www.youtube.com/watch?v=osd-pUSGvlo* | Personal experience | Daith Piercing Video, pain threshold and cleaning. | 25.10.2016 | 5748 | 48 | 388 | Poor | Very poor |
| *90* | *https://www.youtube.com/watch?v=d4yIfdntF4I* | Personal experience | Daith Piercing for Migraine Harley Street - Louisa - Personal Trainer | 31.08.2017 | 5037 | 30 | 405 | Poor | Very poor |
| *91* | *https://www.youtube.com/watch?v=sr0045-4qcU* | Personal experience | Got my daith pierced for migraines. Did it work though? | 27.04.2020 | 3242 | 100 | 850 | Poor | Very poor |
| *92* | *https://www.youtube.com/watch?v=hqSuwG2y18M* | Personal experience | Husband Gets His Daith Pierced \| INSTANT MIGRAINE RELIEF | 09.08.2018 | 3079 | 40 | 424 | Poor | Very poor |
| *93* | *https://www.youtube.com/watch?v=3dLvi8VY12U* | Personal experience | Watch me get a daith piercing for migraines | 20.05.2020 | 2827 | 28 | 482 | Poor | Very poor |
| *94* | *https://www.youtube.com/watch?v=8kDLMkbkWLs* | Personal experience | Migraines suck...so I got my Daith pierced!! | 18.12.2016 | 2097 | 38 | 691 | Poor | Very poor |
| *95* | *https://www.youtube.com/watch?v=K-gHi5ClruU* | Personal experience | Daith Piercing for Migraine Harley Street - Lesley - Mild Blindness | 29.08.2017 | 1734 | 13 | 360 | Poor | Very poor |
| *96* | *https://www.youtube.com/watch?v=4qbyhSr5CA8* | Personal experience | Daith Piercings and Migraines: Part 10 | 09.04.2016 | 703 | 15 | 219 | Poor | Very poor |
| *97* | *https://www.youtube.com/watch?v=kxizmAdLYRo* | Personal experience | Daith Piercing Destroys Migraine using this pre-tested unique method. | 16.12.2021 | 668 | 3 | 115 | Poor | Very poor |
| *98* | *https://www.youtube.com/watch?v=_WhHqXKsnTA* | Personal experience | Daith Piercing for Migraine Sussex - Reena&#39;s Reaction | 08.01.2018 | 514 | 5 | 283 | Poor | Very poor |
| *99* | *https://www.youtube.com/watch?v=At2Z55oXsdw* | Personal experience | Daith Piercing Experience-For Migraines | 26.01.2017 | 365 | 14 | 815 | Poor | Very poor |
| *100* | *https://www.youtube.com/watch?v=HIXHtg-amcE* | Personal experience | MIGRAINE \| DAITH Piercing Update \| Review | 14.12.2020 | 334 | 8 | 378 | Poor | Very poor |
| *101* | *https://www.youtube.com/watch?v=ay4doNSO_c0* | Personal experience | Migraine Dairies \| Can a daith piercing cure migraine? | 08.09.2020 | 330 | 12 | 469 | Poor | Very poor |
| *102* | *https://www.youtube.com/watch?v=yS5OdCWtNqU* | Personal experience | Lisa&#39;s weird Euphoria, &quot;headache gone its weird&quot; - another daith piercing for migraine experience. | 25.06.2021 | 305 | 0 | 157 | Poor | Very poor |
| *103* | *https://www.youtube.com/watch?v=4AEQRiUz8fk* | Personal experience | Fibromyalgia and migraine sufferer Joanne from Swanley “No Headache” from the daith piercing | 08.03.2021 | 267 | 1 | 162 | Poor | Very poor |
| *104* | *https://www.youtube.com/watch?v=z1ZJL8-2Sgk* | Personal experience | Tiana gets the medical daith for head fog, migraine, and anxiety. Explaining what she now feels. | 22.01.2021 | 266 | 1 | 94 | Poor | Very poor |
| *105* | *https://www.youtube.com/watch?v=nqLHSDvUUek* | Personal experience | Daith Piercing For Migraine with Sarah from Croydon at Daith Medical Ltd | 15.03.2021 | 265 | 2 | 168 | Poor | Very poor |
| *106* | *https://www.youtube.com/watch?v=T20J23WnYZA* | Personal experience | Daith Piercing for Migraine Sussex - Katie&#39;s reaction | 04.08.2017 | 261 | 1 | 100 | Poor | Very poor |
| *107* | *https://www.youtube.com/watch?v=sMt8sdKHCOc* | Personal experience | DAITH PIERCING IN MAINE | 15.09.2019 | 258 | 26 | 1020 | Poor | Very poor |
| *108* | *https://www.youtube.com/watch?v=EEtcLCHqpvs* | Personal experience | Daith Piercing Pt 1 | 25.09.2018 | 221 | 1 | 50 | Poor | Very poor |
| *109* | *https://www.youtube.com/watch?v=uDZ7JmKZcRI* | Personal experience | Heather &amp; Andy travel 750 miles for a daith piercing with Daith Medical Ltd, find out if it paid off | 08.10.2020 | 218 | 1 | 180 | Poor | Very poor |
| *110* | *https://www.youtube.com/watch?v=hO_6ZGwLIeQ* | Personal experience | 20yr daily migraine sufferer reduced headaches to nothing with daith | 24.07.2016 | 215 | 6 | 51 | Poor | Very poor |
| *111* | *https://www.youtube.com/watch?v=ubVt_35RUaQ* | Personal experience | VLOG \| I’M GETTING A DAITH PIERCING ( THE HEADACHE/MIGRAINE PIERCING ) \| RIAHJAYSREALITY | 21.01.2021 | 213 | 7 | 1473 | Poor | Very poor |
| *112* | *https://www.youtube.com/watch?v=jZEuIYbqgRQ* | Personal experience | DAITH PIERCING \| Experience + Pain | 13.04.2020 | 211 | 4 | 609 | Poor | Very poor |
| *113* | *https://www.youtube.com/watch?v=6gKvDR6Rcmw* | Personal experience | Daith piercing for migraines 1 yr UPDATE | 11.01.2018 | 208 | 1 | 25 | Poor | Very poor |
| *114* | *https://www.youtube.com/watch?v=RZqPU0iI-Bk* | Personal experience | Daith Piercing for Migraine Harley Street - Katie&#39;s reaction | 04.08.2017 | 184 | 1 | 284 | Poor | Very poor |
| *115* | *https://www.youtube.com/watch?v=uJZQXAtsiag* | Personal experience | Daith Piercing for Migraine Harley Street - | 29.06.2019 | 181 | 1 | 143 | Poor | Very poor |
| *116* | *https://www.youtube.com/watch?v=8kRjCieX9yY* | Personal experience | Migraine Piercing result for young Hannah. Daith Medical Ltd | 08.01.2021 | 173 | 1 | 87 | Poor | Very poor |
| *117* | *https://www.youtube.com/watch?v=mv5oNiGImZ4* | Personal experience | Sickie Girl - Daith Piercings, CBD Oil &amp; Migraines - Comments I&#39;ve Heard. Do They Work? | 05.04.2019 | 152 | 4 | 1552 | Poor | Very poor |
| *118* | *https://www.youtube.com/watch?v=KWNGZQzjjgs* | Personal experience | Daith Piercing for Migraine Sussex - Nurse - Jen | 02.11.2019 | 150 | 1 | 104 | Poor | Very poor |
| *119* | *https://www.youtube.com/watch?v=oecy5Opr3yY* | Personal experience | Daith Piercing Experience- Did it work for my headaches? | 27.12.2016 | 133 | 3 | 294 | Poor | Very poor |
| *120* | *https://www.youtube.com/watch?v=4hw4e4HmOBI* | Personal experience | Daith Piercing for Migraine Harley Street - Jennifer&#39;s Reaction | 17.11.2017 | 106 | 0 | 109 | Poor | Very poor |
| *121* | *https://www.youtube.com/watch?v=vLzaR3eIRkA* | Personal experience | 11. Apr 23 | 11.04.2023 | 94 | 1 | 198 | Poor | Very poor |
| *122* | *https://www.youtube.com/watch?v=kGGygass5aA* | Personal experience | Migraine-Piercing-daithcouk | 14.12.2019 | 90 | 1 | 300 | Poor | Very poor |
| *123* | *https://www.youtube.com/watch?v=ZDLOvuRrl4M* | Personal experience | Daith piercing for migraine Harley Street - Savannah | 02.11.2019 | 74 | 0 | 116 | Poor | Very poor |
| *124* | *https://www.youtube.com/watch?v=FlPIDms-6RU* | Personal experience | Migraine Treatment Joe (full version) Harley Street Daith Piercing | 07.11.2020 | 73 | 0 | 132 | Poor | Very poor |
| *125* | *https://www.youtube.com/watch?v=Uf-aJ8budoE* | Personal experience | Harley-street-daith-piercing | 14.12.2019 | 71 | 2 | 228 | Poor | Very poor |
| *126* | *https://www.youtube.com/watch?v=H-zaH-MgCWc* | Personal experience | Daith Piercing And Migraines | 11.07.2018 | 64 | 2 | 1088 | Poor | Very poor |
| *127* | *https://www.youtube.com/watch?v=zdmiCmGZhWI* | Personal experience | Migraine Relief.. My experience with migraines. | 23.02.2019 | 37 | 0 | 224 | Poor | Very poor |
| *128* | *https://www.youtube.com/watch?v=-rmgWtnuNEo* | Personal experience | Daith Piercing Acupuncture Fusion Therapy | 06.12.2023 | 32 | 0 | 135 | Poor | Very poor |
| *129* | *https://www.youtube.com/watch?v=Lb1I_ZZrJcE* | Personal experience | My migraine story / If the Daith Piercing really helps or not…? (6wk update) | 20.05.2022 | 14 | 2 | 585 | Poor | Very poor |
| *130* | *https://www.youtube.com/watch?v=pZhnjjabi88* | Personal experience | Daith Piercing Infection \| How I Cured It (WITH PICTURES) | 07.10.2016 | 359986 | 4936 | 958 | Very poor | Poor |
| *131* | *https://www.youtube.com/watch?v=nSkE7l8I9zI* | Personal experience | daith piercing to cure migraines ? | 13.05.2018 | 1076 | 22 | 693 | Very poor | Poor |
| *132* | *https://www.youtube.com/watch?v=P0ls4hEKUro* | Personal experience | Changing My Daith Piercing (FIRST TIME)! | 02.03.2018 | 276680 | 3719 | 399 | Very poor | Very poor |
| *133* | *https://www.youtube.com/watch?v=h5G9Wn78yYc* | Personal experience | MOMS DAITH PIERCING EXPIERENCE!! | 03.07.2018 | 71361 |  | 644 | Very poor | Very poor |
| *134* | *https://www.youtube.com/watch?v=1V3WEPIv4S8* | Personal experience | Changing My Daith Jewelry for the First Time | 03.06.2020 | 71288 | 391 | 223 | Very poor | Very poor |
| *135* | *https://www.youtube.com/watch?v=tEtu0hboXEI* | Personal experience | DAITH PIERCING REVIEW! HONEST OPINION... | 11.11.2019 | 8003 | 93 | 490 | Very poor | Very poor |
| *136* | *https://www.youtube.com/watch?v=LU4rAZLnDEY* | Personal experience | Daith piercing and migraines: part 2 | 21.11.2015 | 6488 | 62 | 91 | Very poor | Very poor |
| *137* | *https://www.youtube.com/watch?v=u9DAbrO5EMM* | Personal experience | Getting My Daith Pierced + How Much It Hurts | 25.10.2022 | 5707 | 160 | 90 | Very poor | Very poor |
| *138* | *https://www.youtube.com/watch?v=bRoLlLlwKH0* | Personal experience | Daith piercing and migraines: Part 3 | 28.11.2015 | 5406 | 57 | 134 | Very poor | Very poor |
| *139* | *https://www.youtube.com/watch?v=46yHoN1RtmA* | Personal experience | Daith piercing and migraines: part 4 | 03.12.2015 | 5195 | 56 | 172 | Very poor | Very poor |
| *140* | *https://www.youtube.com/watch?v=EjbeiMY7hms* | Personal experience | Daith piercing and migraines: part 5 | 07.12.2015 | 4676 | 49 | 256 | Very poor | Very poor |
| *141* | *https://www.youtube.com/watch?v=7re80ml3DJs* | Personal experience | Daith Piercing for Migraine Sussex - Jennifer - 9 year sufferer | 06.09.2017 | 4133 | 23 | 340 | Very poor | Very poor |
| *142* | *https://www.youtube.com/watch?v=-Am1wsY_xT4* | Personal experience | MY DAITH EAR PEIRCING &amp; MIGRAINE/HEADCAHE RELIEF | 04.08.2019 | 2085 | 34 | 299 | Very poor | Very poor |
| *143* | *https://www.youtube.com/watch?v=61gzGUcprOQ* | Personal experience | Daith piercing, do they work for migraines? | 27.08.2018 | 1613 | 40 | 770 | Very poor | Very poor |
| *144* | *https://www.youtube.com/watch?v=VW7afcfqjaQ* | Personal experience | Daith piercing and migraines: let&#39;s look at dehydration and supplements | 10.01.2016 | 1603 | 26 | 346 | Very poor | Very poor |
| *145* | *https://www.youtube.com/watch?v=xky4QGFrCMc* | Personal experience | Daith piercings and migraines: part 12 | 29.05.2016 | 1406 | 23 | 279 | Very poor | Very poor |
| *146* | *https://www.youtube.com/watch?v=D5DnQ4C8cLA* | Personal experience | Daith Piercing for Migraine Sussex - Emma | 02.11.2019 | 1282 | 5 | 119 | Very poor | Very poor |
| *147* | *https://www.youtube.com/watch?v=pFpXnoMA7zQ* | Personal experience | daith piercing and psoriasis update | 31.05.2018 | 1152 | 14 | 642 | Very poor | Very poor |
| *148* | *https://www.youtube.com/watch?v=_i1IbH566jA* | Personal experience | Daith Piercing Cures Migraines?! | 15.12.2017 | 1040 | 20 | 276 | Very poor | Very poor |
| *149* | *https://www.youtube.com/watch?v=w2jxDLP5DjE* | Personal experience | Daith Piercing for Migraine Sussex - Results | 06.04.2017 | 1039 | 9 | 56 | Very poor | Very poor |
| *150* | *https://www.youtube.com/watch?v=bXsgqEHveTQ* | Personal experience | #shorts Migraine Daith ear piercing update👂 | 26.04.2023 | 593 | 41 | 48 | Very poor | Very poor |
| *151* | *https://www.youtube.com/watch?v=p_BCFEqkL5c* | Personal experience | I GOT MY DAITH PIERCED TO STOP MIGRAINES/MY EXPERIENCE | 09.02.2020 | 523 | 25 | 531 | Very poor | Very poor |
| *152* | *https://www.youtube.com/watch?v=1IUvGTXko5U* | Personal experience | Daith Piercing for Migraines | 09.12.2019 | 491 | 8 | 121 | Very poor | Very poor |
| *153* | *https://www.youtube.com/watch?v=E857Vvtv9AQ* | Personal experience | Daith Piercing for Migraine Sussex - Raimondas - Window Cleaner | 02.11.2018 | 440 | 2 | 100 | Very poor | Very poor |
| *154* | *https://www.youtube.com/watch?v=HQL72LGfg-8* | Personal experience | Influencer Mariane Daith Piercing For Migraine at Daith Medical Ltd Burgess Hill Studio | 03.12.2020 | 429 | 7 | 110 | Very poor | Very poor |
| *155* | *https://www.youtube.com/watch?v=am800FHeW0w* | Personal experience | Migraine relief - getting the daith piercing | 20.02.2019 | 429 | 5 | 668 | Very poor | Very poor |
| *156* | *https://www.youtube.com/watch?v=YImpfkg1lsQ* | Personal experience | Daith Piercing for migraine Sussex - Katherine - Beautician | 02.11.2019 | 375 | 0 | 229 | Very poor | Very poor |
| *157* | *https://www.youtube.com/watch?v=yiki9kzGyio* | Personal experience |  | 10.05.2022 | 333 | 5 | 259 | Very poor | Very poor |
| *158* | *https://www.youtube.com/watch?v=CRFOMpBAMb0* | Personal experience | Daith Piercing for Migraine Sussex - Nigel - Double Migraine Cured with VNS | 29.11.2017 | 330 | 2 | 151 | Very poor | Very poor |
| *159* | *https://www.youtube.com/watch?v=2jUM68Gqgi0* | Personal experience | Daith Piercing for Migraine Sussex - Jane - 10 Year sufferer from fall at work | 23.01.2018 | 327 | 4 | 299 | Very poor | Very poor |
| *160* | *https://www.youtube.com/watch?v=wQ9t5vvVZHA* | Personal experience | Daith Piercing for Migraine Sussex - Hannah&#39;s Reaction | 08.01.2018 | 320 | 0 | 197 | Very poor | Very poor |
| *161* | *https://www.youtube.com/watch?v=kaQkU_YsHWs* | Personal experience | Does a daith piercing cure migraines? Vlog #10 | 12.08.2016 | 238 | 2 | 362 | Very poor | Very poor |
| *162* | *https://www.youtube.com/watch?v=oiRVjHxbkyg* | Personal experience | Midwife Laura gets a daith piercing for migraine and anxiety. | 21.12.2019 | 238 | 1 | 118 | Very poor | Very poor |
| *163* | *https://www.youtube.com/watch?v=r1GKfTW0Uow* | Personal experience | Will gets the Daith Piercing for Migraine/Tension Headache from Daith Medical Ltd | 24.12.2020 | 232 | 0 | 57 | Very poor | Very poor |
| *164* | *https://www.youtube.com/watch?v=yxbgZ_0iWG0* | Personal experience | Migraine sufferer Holly from Leigh on Sea get the pre-tested medical daith piercing | 15.04.2021 | 226 | 6 | 162 | Very poor | Very poor |
| *165* | *https://www.youtube.com/watch?v=oVUI-XDOQ18* | Personal experience | Migraine Piercing (pre-tested) Daith Piercing Testimonial from Trish | 18.11.2020 | 205 | 1 | 233 | Very poor | Very poor |
| *166* | *https://www.youtube.com/watch?v=d8LDhoEuvQQ* | Personal experience | Daith Piercing | 22.12.2019 | 200 |  | 28 | Very poor | Very poor |
| *167* | *https://www.youtube.com/watch?v=JEGslQqKGT4* | Personal experience | Daith Piercing for Migraines? | 21.07.2018 | 185 | 2 | 153 | Very poor | Very poor |
| *168* | *https://www.youtube.com/watch?v=nXkP5RyVwyI* | Personal experience | Daith Piercing for Migraine Sussex - Nicola - Migraines since age 9 | 16.01.2018 | 177 | 1 | 150 | Very poor | Very poor |
| *169* | *https://www.youtube.com/watch?v=S3lDqOruvW4* | Personal experience | Daith Piercing For Migraine | 14.12.2019 | 173 | 2 | 199 | Very poor | Very poor |
| *170* | *https://www.youtube.com/watch?v=0R2cXohCmDk* | Personal experience | Daith Piercings Update -- 4 Months -- Daiths Vs. Migraines | 16.02.2017 | 159 | 2 | 292 | Very poor | Very poor |
| *171* | *https://www.youtube.com/watch?v=essVZ7dTRN8* | Personal experience | Daith piercing | 15.09.2017 | 127 | 2 | 86 | Very poor | Very poor |
| *172* | *https://www.youtube.com/watch?v=MkYEo5RqqOo* | Personal experience | THE MIGRAINE DAITH PIERCING \| got my daith pierced | 30.09.2019 | 120 | 2 | 314 | Very poor | Very poor |
| *173* | *https://www.youtube.com/watch?v=f1UCZ7vvxDQ* | Personal experience | Daith/migraine and infection! | 06.12.2021 | 117 | 1 | 16 | Very poor | Very poor |
| *174* | *https://www.youtube.com/watch?v=ql0PNdZ4ncE* | Personal experience | Daith Piercings Vs. Migraines | 10.12.2016 | 111 | 1 | 441 | Very poor | Very poor |
| *175* | *https://www.youtube.com/watch?v=PCAT2wNFrYs* | Personal experience | Daith Piercing for Migraines - Smile! | 11.04.2020 | 81 | 3 | 232 | Very poor | Very poor |
| *176* | *https://www.youtube.com/watch?v=xa_FjneqILU* | Personal experience | #daith #piercing #questions #migraine #infections Please watch other video for more information | 21.04.2023 | 56 | 2 | 60 | Very poor | Very poor |
| *177* | *https://www.youtube.com/watch?v=vD7Fn55dnfs* | Personal experience | No more Migraines!!!( Daith Piercing ) | 05.02.2023 | 54 | 4 | 320 | Very poor | Very poor |
| *178* | *https://www.youtube.com/watch?v=hrcajy_l4Ho* | Others | Fad Friday: Daith Piercings for Migraines | 19.02.2016 | 2889 | 13 | 168 | Excellent | Good |
| *179* | *https://www.youtube.com/watch?v=hFoHLMWyHd8* | Others | Daith piercing and migraine correlation explanation by acupuncture specialist | 06.08.2017 | 541 | 6 | 128 | Excellent | Good |
| *180* | *https://www.youtube.com/watch?v=putikpTx9t8* | Others | What You Should Know About Daith Ear Piercings For Migraines | 03.05.2020 | 84 | 0 | 162 | Excellent | Good |
| *181* | *https://www.youtube.com/watch?v=wJ-Cid7S8A0* | Others | 🟡 Daith Piercing For Migraine And Headache Relief? | 22.07.2021 | 48 | 0 | 220 | Excellent | Good |
| *182* | *https://www.youtube.com/watch?v=L9FfXB_G-SE* | Others | Daith Ear Piercing for Migraines \| Headache Myth or Miracle? | 03.05.2019 | 2638 | 29 | 193 | Excellent | Moderate |
| *183* | *https://www.youtube.com/watch?v=b8WLdWi3BzI* | Others | Piercing Realtalk Episode 4 : The Truth About Daiths and Migraines | 23.12.2017 | 4979 | 235 | 663 | Good | Good |
| *184* | *https://www.youtube.com/watch?v=GtKjX24_BCg* | Others | Piercer Discusses Migraine Cure Piercing | 18.11.2018 | 75118 | 1696 | 414 | Good | Moderate |
| *185* | *https://www.youtube.com/watch?v=epOzK4G0tg0* | Others | Ear piercing may stop sever migraines | 05.05.2018 | 271 | 2 | 217 | Good | Moderate |
| *186* | *https://www.youtube.com/watch?v=2izHd7TG7oQ* | Others | Can tragus or daith piercings help with migraine relief? | 18.07.2023 | 176 | 5 | 87 | Good | Moderate |
| *187* | *https://www.youtube.com/watch?v=E1N_cn-hcuA* | Others | Daith piercing: Could it solve your migraine problem? | 04.11.2016 | 86491 | 561 | 285 | Good | Poor |
| *188* | *https://www.youtube.com/watch?v=fqNeTtFyPtE* | Others | DAITH PIERCING &amp; MIGRAINES | 15.04.2019 | 389 | 4 | 231 | Good | Poor |
| *189* | *https://www.youtube.com/watch?v=WxI8F4voCcA* | Others | Live Eng 12 - Daith and Migraines | 10.06.2022 | 7 | 0 | 736 | Good | Poor |
| *190* | *https://www.youtube.com/watch?v=uCWOcgxoMT4* | Others | Do Daith Piercings Cure Migraines?- THE MODIFIED WORLD | 30.11.2015 | 23932 | 1123 | 324 | Fair | Good |
| *191* | *https://www.youtube.com/watch?v=SGoYpJxsSlo* | Others | Daith Piercing Pros &amp; Cons by a Piercer EP 01 | 15.01.2019 | 50911 | 1331 | 713 | Fair | Moderate |
| *192* | *https://www.youtube.com/watch?v=n-dKvdvhbzc* | Others | Do “Daith” Piercings Help Migraines? | 18.02.2016 | 10276 | 35 | 93 | Fair | Moderate |
| *193* | *https://www.youtube.com/watch?v=mXzM-NpcQ9c* | Others | The Facts: Do Daith Piercings Cure Migraines? - The Truth About Daith Piercings | 21.07.2017 | 805 | 10 | 317 | Fair | Moderate |
| *194* | *https://www.youtube.com/watch?v=jooDO4RNJ8I* | Others | This Piercing Cures Migraines? | 21.02.2021 | 9060 | 389 | 199 | Fair | Poor |
| *195* | *https://www.youtube.com/watch?v=CEMWa4l-kXI* | Others | Migraine, Headache Miracle Solutions, Piercings &amp; Acupuncture with Dr. Ken Grey on ABC News | 25.05.2017 | 1151 | 11 | 257 | Fair | Poor |
| *196* | *https://www.youtube.com/watch?v=eW9hQcR8u0k* | Others | People continue turning to piercing for headache relief | 08.05.2019 | 385 | 2 | 79 | Fair | Poor |
| *197* | *https://www.youtube.com/watch?v=0-_sqAGgmtg* | Others | KSL Migraine Miracle | 17.05.2017 | 277 | 1 | 258 | Fair | Poor |
| *198* | *https://www.youtube.com/watch?v=JhyCZcscVYA* | Others | Dothan piercing parlor pierces migraine pain away | 12.07.2017 | 137 | 0 | 195 | Fair | Poor |
| *199* | *https://www.youtube.com/watch?v=DccxHQN18g4* | Others | Are Tragus And Daith Piercings Curing Migraines? Video | 14.04.2019 | 4630 | 66 | 629 | Poor | Moderate |
| *200* | *https://www.youtube.com/watch?v=Ndx_6Q-P7pc* | Others | Daith Piercing Info &amp; Aftercare \| UrbanBodyJewelry.com | 06.11.2017 | 187812 | 4194 | 310 | Poor | Poor |
| *201* | *https://www.youtube.com/watch?v=G5QLGP1Tl8E* | Others | 5 Things We HATE About Daith Piercings!! 🤬 | 19.06.2020 | 91963 |  | 282 | Poor | Poor |
| *202* | *https://www.youtube.com/watch?v=FzHv71JuzsA* | Others | Can a Body Piercing Stop Migraines? | 27.02.2016 | 44657 | 218 | 187 | Poor | Poor |
| *203* | *https://www.youtube.com/watch?v=5yGfi477K54* | Others | Daith Piercing What You Should Know Consultations by a Piercer EP12 | 15.08.2022 | 4769 | 143 | 778 | Poor | Poor |
| *204* | *https://www.youtube.com/watch?v=wHlARtJzPf0* | Others | Le piercing thérapeutique pour lutter contre les migraines ? | 27.07.2020 | 1562 | 32 | 568 | Poor | Poor |
| *205* | *https://www.youtube.com/watch?v=KSmm02VGudY* | Others | Daith piercings to treat migraines | 28.01.2020 | 777 | 4 | 174 | Poor | Poor |
| *206* | *https://www.youtube.com/watch?v=9P72_hycu18* | Others | Piercings for Migraine Relief: Do daith &amp; tragus piercings really help?? #shorts | 18.07.2023 | 717 | 15 | 60 | Poor | Poor |
| *207* | *https://www.youtube.com/watch?v=x99gZ8lI_ME* | Others | London Migraine Clinic - First evidence that Daith Piercing for Migraines has a medical effect | 05.12.2021 | 434 | 7 | 947 | Poor | Poor |
| *208* | *https://www.youtube.com/watch?v=vTS32sXwrLg* | Others | ALEXIS DEL CID KCTV5 responds to Daith Piercing story comments | 16.11.2016 | 272 | 4 | 91 | Poor | Poor |
| *209* | *https://www.youtube.com/watch?v=eZvH9FGKvgc* | Others | Daith Piercing Update; Migraine Theory. | 28.10.2016 | 46 | 4 | 484 | Poor | Poor |
| *210* | *https://www.youtube.com/watch?v=qWWHt4bM9AM* | Others | Daith Piercing as A Cure to Migraine? Acupuncturists &amp; Piercing Aftercare | 22.11.2022 | 11 | 0 | 86 | Poor | Poor |
| *211* | *https://www.youtube.com/watch?v=G9BY5_AFA_w* | Others | Alexis Del Cid, KCTV5 Investigates Daith Piercing | 04.11.2016 | 69966 | 292 | 199 | Poor | Very poor |
| *212* | *https://www.youtube.com/watch?v=eXlqIxKdRJI* | Others | ASMR Daith Piercing RolePlay For Migraines \| Soft Spoken | 10.08.2017 | 16180 | 378 | 1029 | Poor | Very poor |
| *213* | *https://www.youtube.com/watch?v=s7csgFam2Ak* | Others | Why we HATE daith piercings #shorts | 02.08.2022 | 14595 | 405 | 50 | Poor | Very poor |
| *214* | *https://www.youtube.com/watch?v=DR3pOL3VBTQ* | Others | Daith Piercing Jewelry Pros &amp; Cons by a Piercer EP 56 | 16.02.2020 | 5934 | 230 | 913 | Poor | Very poor |
| *215* | *https://www.youtube.com/watch?v=UL5y9BveO2E* | Others | Could this piercing help stop your migraines? | 09.06.2017 | 1182 | 7 | 183 | Poor | Very poor |
| *216* | *https://www.youtube.com/watch?v=F851SBh562g* | Others | Daith Piercing for Migraine | 27.07.2022 | 561 | 3 | 59 | Poor | Very poor |
| *217* | *https://www.youtube.com/watch?v=h6ldxUCTLd0* | Others | People continue turning to &#39;daith&#39; piercing for headache relief | 02.05.2019 | 376 | 1 | 101 | Very poor | Poor |
| *218* | *https://www.youtube.com/watch?v=TK-WwJOaUd8* | Others | Daith Piercing (migraine/headache piercing) | 12.12.2019 | 116777 | 2556 | 179 | Very poor | Very poor |
| *219* | *https://www.youtube.com/watch?v=4D8mombpsfA* | Others | Daith Piercing for Migraine Harley Street - Lindsey&#39;s Reaction | 17.11.2017 | 70134 | 10 | 221 | Very poor | Very poor |
| *220* | *https://www.youtube.com/watch?v=YnESzhQlX8A* | Others | Daith Piercing Pros &amp; Cons \| Lulu&#39;s Body Piercing | 03.10.2019 | 33574 |  | 132 | Very poor | Very poor |
| *221* | *https://www.youtube.com/watch?v=1TlQFvGMwy0* | Others | When is a Daith Piercing Healed? \| UrbanBodyJewelry.com | 28.05.2021 | 7340 | 141 | 122 | Very poor | Very poor |
| *222* | *https://www.youtube.com/watch?v=azQCZ8oTHvI* | Others | The Daith Piercing: Possible Migraine Remedy | 12.06.2017 | 6755 | 45 | 201 | Very poor | Very poor |
| *223* | *https://www.youtube.com/watch?v=N5NweFRvypw* | Others | daith piercing migraine relief instantly | 21.08.2017 | 4195 | 40 | 142 | Very poor | Very poor |
| *224* | *https://www.youtube.com/watch?v=88U3AWmtoBw* | Others | Does DAITH PIERCING help in treating MIGRAINE? | 09.11.2022 | 2569 | 65 | 56 | Very poor | Very poor |
| *225* | *https://www.youtube.com/watch?v=5gGFyd5Ba9Y* | Others | Are daith piercings a cure for headaches? | 12.12.2015 | 1079 | 4 | 184 | Very poor | Very poor |
| *226* | *https://www.youtube.com/watch?v=IxhYnjF6cFI* | Others | Daith Piercing for Migraine Sussex - Joanne - Migraines Since Age 8 | 25.04.2018 | 750 | 7 | 213 | Very poor | Very poor |
| *227* | *https://www.youtube.com/watch?v=FYoOapGur-w* | Others | 8 years daith piercing comes out with instant migraine coming back. | 05.05.2021 | 602 | 5 | 175 | Very poor | Very poor |
| *228* | *https://www.youtube.com/watch?v=Y1dwxK4Osmk* | Others | Daith Piercing | 17.11.2017 | 415 | 0 | 102 | Very poor | Very poor |
| *229* | *https://www.youtube.com/watch?v=F0zYCT-RFGQ* | Others | People continue turning to &#39;daith&#39; piercing for headache relief | 02.05.2019 | 320 | 3 | 107 | Very poor | Very poor |
| *230* | *https://www.youtube.com/watch?v=ecEZL0SO_58* | Others | Cheektowaga tattoo shop has remedy for chronic migraines | 29.04.2017 | 315 | 1 | 198 | Very poor | Very poor |
| *231* | *https://www.youtube.com/watch?v=h0ej5ne8w1w* | Others | DAITH PIERCING for DEBILITATING &amp; PAINFUL MIGRAINES | 20.05.2022 | 292 | 11 | 127 | Very poor | Very poor |
| *232* | *https://www.youtube.com/watch?v=z2ntkHD4v8g* | Others | Daith Piercing for Migraine Harley Street - Christine&#39;s Reaction | 29.11.2017 | 286 | 1 | 160 | Very poor | Very poor |
| *233* | *https://www.youtube.com/watch?v=YOhWOVBxH2w* | Others | Daith Medical Ltd CEO Richard Soper introducing his You Tube Channel | 24.12.2020 | 241 | 2 | 36 | Very poor | Very poor |
| *234* | *https://www.youtube.com/watch?v=tllVZk30Zks* | Others | Daith Piercing for Migraine Sussex - Jo&#39;s Reaction | 17.11.2017 | 191 | 0 | 268 | Very poor | Very poor |
| *235* | *https://www.youtube.com/watch?v=l-HfJjgkOYQ* | Others | Daith re-piercing after 3 years. Caroline from Horsham feeling so much better straight away. | 02.11.2022 | 191 | 3 | 132 | Very poor | Very poor |
| *236* | *https://www.youtube.com/watch?v=4mSf-6US7t4* | Others | Daith Piercing for Migraine Sussex - Dave - 7 Months On | 15.01.2018 | 185 | 1 | 91 | Very poor | Very poor |
| *237* | *https://www.youtube.com/watch?v=exad5IaErlY* | Others | Daith Piercing for Migraine Harley Street - Charlotte&#39;s Reaction | 17.11.2017 | 158 | 0 | 131 | Very poor | Very poor |
| *238* | *https://www.youtube.com/watch?v=psYJSNOXJ50* | Others | Daith Piercing for Migraine Harley Street - Rachiel&#39;s Reaction | 17.11.2017 | 154 | 0 | 84 | Very poor | Very poor |
| *239* | *https://www.youtube.com/watch?v=Kv2ZgEDAu3U* | Others | Daith Piercing for Migraine Sussex - Jodie - 16 year Migraine | 02.11.2019 | 129 | 1 | 167 | Very poor | Very poor |
| *240* | *https://www.youtube.com/watch?v=hx93XZTn5QM* | Others | Instant Migraine relief for fibromyalgia sufferer Nat and several of her family | 08.02.2020 | 125 | 2 | 235 | Very poor | Very poor |
| *241* | *https://www.youtube.com/watch?v=cwtv9a4AFu8* | Others | DaithPiercing | 23.11.2019 | 124 | 2 | 196 | Very poor | Very poor |
| *242* | *https://www.youtube.com/watch?v=v033OxFdp3U* | Others | Daith Piercing for Migraine Harley Street - Louise&#39;s Reaction | 17.11.2017 | 108 | 0 | 230 | Very poor | Very poor |
| *243* | *https://www.youtube.com/watch?v=7YnlZp6Df04* | Others | Harley Street Daith Piercing for Migraine | 13.11.2019 | 87 | 0 | 182 | Very poor | Very poor |
| *244* | *https://www.youtube.com/watch?v=pKWyMCpTlOs* | Others | Migraine Piercing for Gary (Business Consultant) with Researcher and Consultant Richard Soper | 03.12.2020 | 86 | 0 | 88 | Very poor | Very poor |
| *245* | *https://www.youtube.com/watch?v=Aau_WdpfQww* | Others | Migraine gone with just one needle | 29.09.2023 | 48 | 2 | 111 | Very poor | Very poor |
| *246* | *https://www.youtube.com/watch?v=nGNUTKCMnyk* | Others | Daith Piercing for Migraine Sussex - Adreanne - Compounded Headaches | 14.12.2017 | 25 | 0 | 107 | Very poor | Very poor |
